# Saliva viral load is a dynamic unifying correlate of COVID-19 severity and mortality

**DOI:** 10.1101/2021.01.04.21249236

**Authors:** Julio Silva, Carolina Lucas, Maria Sundaram, Benjamin Israelow, Patrick Wong, Jon Klein, Maria Tokuyama, Peiwen Lu, Arvind Venkataraman, Feimei Liu, Tianyang Mao, Ji Eun Oh, Annsea Park, Arnau Casanovas-Massana, Chantal B. F. Vogels, M. Catherine Muenker, Joseph Zell, John B. Fournier, Melissa Campbell, Michael Chiorazzi, Edwin Ruiz Fuentes, Mary E Petrone, Chaney C. Kalinich, Isabel M. Ott, Annie Watkins, Adam J. Moore, Maura Nakahata, Yale IMPACT Team, Nathan D. Grubaugh, Shelli Farhadian, Charles Dela Cruz, Albert I. Ko, Wade L. Schulz, Aaron Ring, Shuangge Ma, Saad Omer, Anne L Wyllie, Akiko Iwasaki

**Author notes:** **Correspondence and requests for materials** should be addressed to A.I. A list of members and their affiliations appears at the end of the paper (Yale IMPACT Team). Lead contact: A.I.

## Abstract

While several clinical and immunological parameters correlate with disease severity and mortality in SARS-CoV-2 infection, work remains in identifying unifying correlates of coronavirus disease 2019 (COVID-19) that can be used to guide clinical practice. Here, we examine saliva and nasopharyngeal (NP) viral load over time and correlate them with patient demographics, and cellular and immune profiling. We found that saliva viral load was significantly higher in those with COVID-19 risk factors; that it correlated with increasing levels of disease severity and showed a superior ability over nasopharyngeal viral load as a predictor of mortality over time (AUC=0.90). A comprehensive analysis of immune factors and cell subsets revealed strong predictors of high and low saliva viral load, which were associated with increased disease severity or better overall outcomes, respectively. Saliva viral load was positively associated with many known COVID-19 inflammatory markers such as IL-6, IL-18, IL-10, and CXCL10, as well as type 1 immune response cytokines. Higher saliva viral loads strongly correlated with the progressive depletion of platelets, lymphocytes, and effector T cell subsets including circulating follicular CD4 T cells (cTfh). Anti-spike (S) and anti-receptor binding domain (RBD) IgG levels were negatively correlated with saliva viral load showing a strong temporal association that could help distinguish severity and mortality in COVID-19. Finally, patients with fatal COVID-19 exhibited higher viral loads, which correlated with the depletion of cTfh cells, and lower production of anti-RBD and anti-S IgG levels. Together these results demonstrated that viral load – as measured by saliva but not nasopharyngeal — is a dynamic unifying correlate of disease presentation, severity, and mortality over time.

---

COVID-19 is a disease caused by the novel SARS-CoV-2 virus, which results in a heterogenous presentation of symptoms and severity. While most cases of COVID-19 are asymptomatic and mild, the full spectrum of disease includes more serious infections ranging from moderate to severe, requiring increasing levels of medical intervention, and may ultimately prove fatal. Though patients with severe disease have been reported in all spectrums of age and health, males, older patients, and those with specific underlying medical conditions are associated with increased disease severity, while those with multiple of these COVID-19 health risk factors are associated with an even greater risk of severe disease^1-4^.

Clinically, severe COVID-19 is characterized by the induction of a strong inflammatory immune response, lymphopenia, thrombocytopenia and coagulopathies^5-7^. Patients with severe disease that progress to death have been shown to express higher levels of inflammatory cytokines and chemokines (CXCL10, IL-6, IL-10), the inflammasome (IL-18, IL-1β), and interferons (IFNα, IFNγ, IFNλ), as well as the induction of type II and III immune responses^8-10^. Furthermore, studies have revealed the benefit of cellular and humoral adaptive immune responses for improved disease outcomes^11-13^, as well as the reduction in follicular CD4 T cell subsets needed for the development and maturation of high-quality antibodies^14^. Despite these associations, to date no factor has been described that can significantly distinguish the full spectrum of severity in COVID-19, from mild to fatal manifestations of disease as well as unify the many cellular, immunological, clinical, and demographic parameters seen in the spectrum of disease severity.

The study of viral load in COVID-19, obtained from distinct collection sites including nasopharynx, sputum, saliva, plasma, urine, and stool, have demonstrated that the information obtained from each of these sites is not always synonymous^15-29^. The association of nasopharyngeal viral load to disease severity has been the most studied. While some studies have shown that high nasopharyngeal viral loads on admission are significantly associated with a higher risk of mortality, older age, and some COVID-19 health risk factors, and others have shown a steady decline of nasopharyngeal viral load amongst those hospitalized with moderate disease, other studies have not found these associations^8,15,27,28,30-32^. Nasopharyngeal viral load has been shown to positively correlate with important inflammatory factors seen in SARS-CoV-2 including TNFα and type I and II interferons, and plasma viremia, which has also been associated with disease severity, to lymphopenia^8,33^.

Saliva viral load in SARS-CoV-2 has been shown to be a viable alternative to the detection of COVID-19 ^29,34^. Similar to some nasopharyngeal studies, viral load obtained from posterior oropharyngeal saliva samples, a distinct sample collection site from standard saliva samples, have been positively associated with age^35^. Given the associations that nasopharyngeal viral load has on specific aspects of COVID-19 disease, our work sought to study the associations that saliva viral load had on important parameters of COVID-19 including the association with known risk factors, and hallmark disease phenotypes such as strong cytokine induction, lymphopenia, and its association with immature humoral immune responses.

## Summary of Cohort Composition

One hundred and fifty four patients admitted to Yale-New Haven hospital between March and June of 2020 were used for analysis as part of our ongoing longitudinal IMPACT study^8^. We assessed nasopharyngeal and saliva viral RNA load quantified by RT-qPCR, as well as levels of plasma cytokines, chemokines and antibodies. Leukocyte profiles were obtained from freshly isolated peripheral blood mononuclear cells (PBMCs). In addition, clinical data was obtained from patient healthcare records and were matched to collection timepoints within a maximum of 48 hours difference from the date of saliva or nasopharyngeal sample collection. Our cohort was comprised of different subgroups categorized by gradations of disease severity including 62 hospitalized patients who developed severe disease and were admitted to the ICU during their hospital stay, and 84 hospitalized patients with moderate disease symptoms. A total of 23 patients with COVID-19 died during the time of our study. A separate cohort of individuals not hospitalized for COVID-19, called “non-hospitalized” in our study, consisted of healthcare workers who were being serially monitored for infection and who became RT-qPCR positive by nasopharyngeal swab or saliva during the study but did not require hospitalization for symptoms. In addition, patients who were admitted for elective care and were discovered to be positive via nasopharyngeal swab RT-qPCR screening but who did not require medical intervention for their COVID-19 infection throughout their stay were also included. In total these groups included 18 RT-qPCR positive healthcare workers who were not hospitalized for symptoms and 8 RT-qPCR positive patients who were not hospitalized for COVID-19. Finally, 108 uninfected healthcare workers served as a control group. A comprehensive demographic breakdown of the cohort is provided in Extended Data Table 1. A total of 355 samples were collected from the infected cohort, which included 1-8 collection timepoints from participants spanning from 2 days before until 40 days after the onset of symptoms. To gain a better understanding of the relationship between viral load and the parameters measured, individuals with measured saliva viral loads were stratified into three groups based on the quantile distribution of viral load levels across the entire cohort, as low (saliva viral load=3.212-4.4031 Log_10_ [Log](GE/ml)), medium (saliva viral load=4.4031-6.1106 Log (GE/ml)), and high (saliva viral load=6.1106-10.320 Log (GE/ml)), irrespective of disease severity for all timepoints collected. Our threshold viral load limit of detection was 3.22 Log (GE/ml) labeled in this study as viral load limit of detection (VLD), and our threshold limit of positivity, labeled as viral load limit of positivity (VLP) was 3.75 Log (GE/ml).

### First-recorded saliva viral load associates with COVID-19 risk factors

Male sex, older age, and specific respiratory, cardiovascular, oncological, and other systemic and immune-suppressive conditions are associated with worse outcomes in COVID-19 ^1-4^. To assess if viral load may be linked with these risk factors, we analyzed first-recorded saliva and nasopharyngeal viral load levels against patient demographics known to be associated with worse outcomes. Samples collected within two weeks following the administration of convalescent serum were excluded from this analysis. Patients were categorized based on the presence or absence of a particular COVID-19 health risk factor (Extended Table 1), which included chronic respiratory conditions (e.g., COPD and asthma), as well as cardiovascular disease, hypertension, cancer within one year of infection, and other conditions that lead to immunosuppression including HIV infection, type II diabetes, cirrhosis, chronic kidney disease, or being actively treated with a drug known to cause immunosuppression. Saliva viral load was significantly higher in patients who were found to have any of these five categorical health risk factors (Extended Figure 1). To assess if these effects were additive, given that individuals with multiple health risk factors are associated with a higher risk of severe disease^4^, patients were then stratified into five categories representing their summative risk factors from 0 to 4+. These categories demonstrated a significant increasing positive trend of disease severity and mortality (Extended Figure 1c, d). Notably, COVID-19 health risk factors demonstrated significant increasing measurements of saliva viral load in which patients with four or more risk factors had significantly higher saliva viral loads than those with zero, or one, or two risk factors upon first sample collection (Figure 1a). Age also showed a significant positive correlation with saliva viral load (Figure 1b) and comparison of viral load between sex demonstrated that saliva viral load was significantly higher in male than in female patients (Figure 1c). By contrast nasopharyngeal viral load was not statistically significantly related to any of these parameters that were measured (Figure 1d, e, f and Extended Figure 1 f-j). These results suggest that risk factors for COVID-19 may be reflective of an underlying replicative capacity of the virus found in the saliva of these individuals from the first collection that appears additive and is associated with an increased likelihood of severity and mortality.

**Fig. 1.**
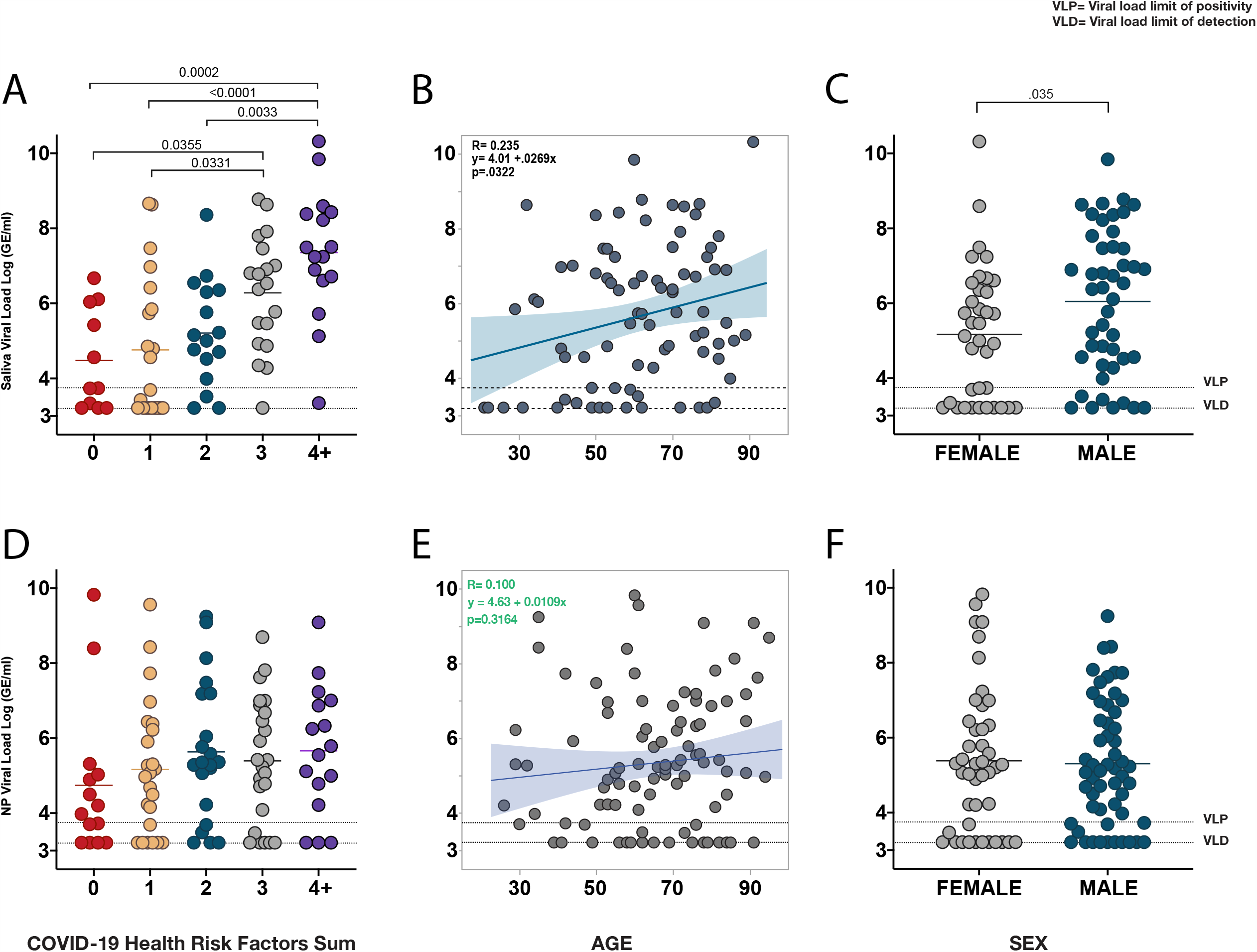
Saliva viral load is significantly higher in individuals with multiple health risk factors, older patients, and in males. **a-f, a**, Comparison of saliva viral load amongst patients stratified from 0 to 4+, representing their cumulative pre-existing COVID-19 health risk factors (Extended Figure 1 and Table 1). **b**, Linear Regression of age and saliva viral load. c, t-test comparing saliva viral load by sex. **d**,**e**,**f**, Comparison of nasopharyngeal (NP) viral load to factors described in **a, b**, and **c**, respectively. All data points represent the first-recorded viral load measurement of a patient for saliva or nasopharyngeal viral load. Solid horizontal lines represent the mean. Significance of comparison for **a** and **c**, was determined by one-way ANOVA followed by Tukey’s test to account for multiple comparisons. Linear regression shows Pearson’s correlation coefficients; shading represents the 95% confidence interval for the regression line. Dotted lines VLP=positivity threshold for viral load is 3.75 Log (Genomic Equivalents [GE]/ml)); VLD=viral load limit of detection is 3.22 Log (GE/ml).

### Early saliva viral load correlates with a spectrum of disease severity and mortality

We then sought to compare viral load with respect to the spectrum of disease severity in our cohort. To do so, we first compared viral load measurements between hospitalized and non-hospitalized individuals (described above) using only first-recorded viral load within the first ten days from symptom onset in order to more closely match stage of disease. Hospitalized patients demonstrated significantly higher saliva viral loads than non-hospitalized individuals (Figure 2a). Comparison of first recorded saliva viral loads for hospitalized patients between moderate and severe disease and alive and deceased individuals throughout the course of disease revealed that saliva viral load was progressively and significantly higher in patients with worse outcomes (Figure 2b, c).

**Fig. 2.**
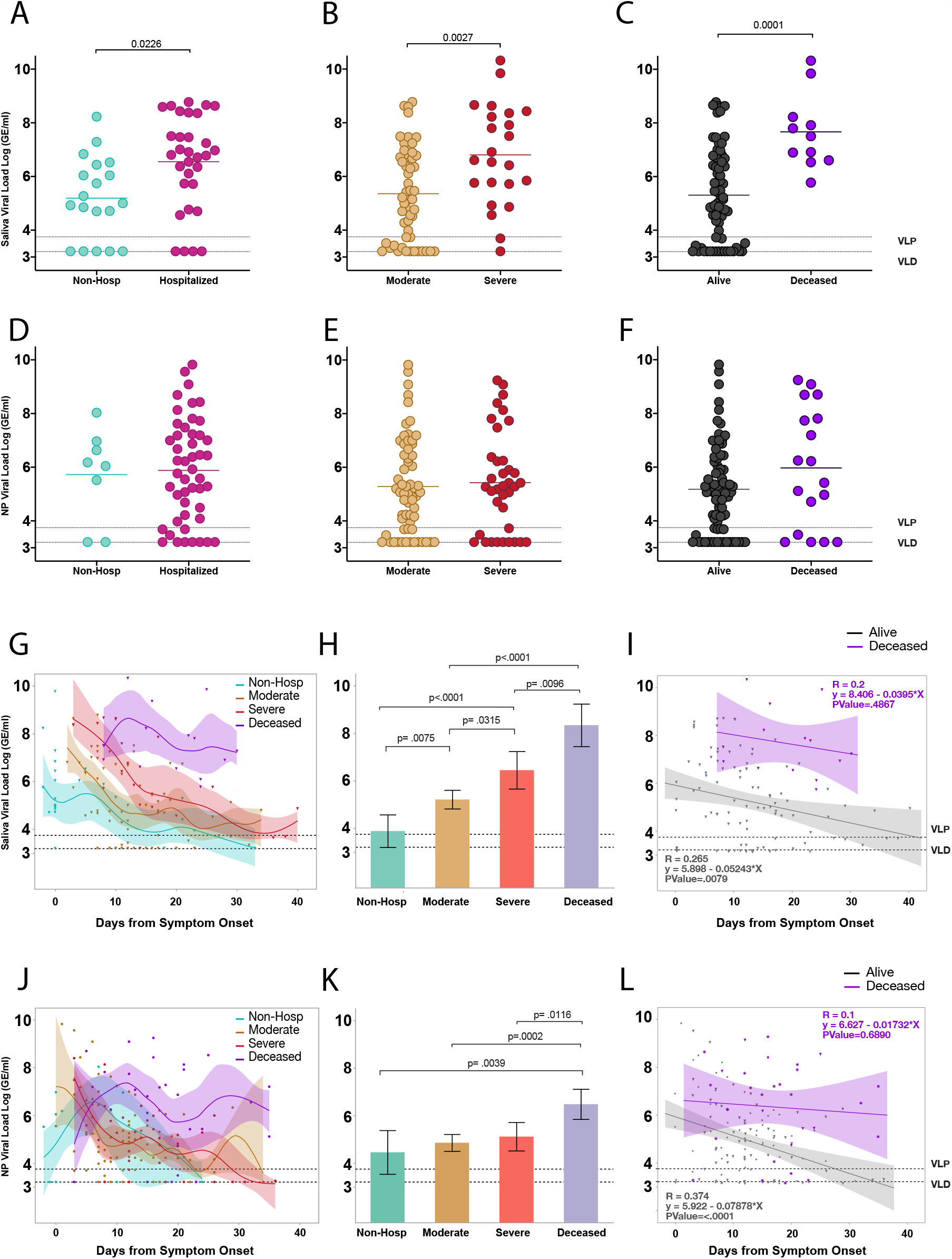
Saliva viral load is strongly associated with spectrum of disease severity throughout the course of illness. **a-l, a**, Comparison of first recorded saliva viral load between individuals hospitalized for COVID-19 and non-hospitalized individuals within the first 10 days from symptom onset using a two-sided t-test. Comparison of only first recorded saliva viral load measurements amongst (b) moderate and severe disease or (c) alive and deceased individuals throughout the course of disease. **d, e, f**, comparison of nasopharyngeal viral load as in **a, b**, and **c** respectively. Significance was adjusted for multiple comparisons within viral load groups using the Holm-Sidak method. **g**, Comparison of saliva viral load between non-hospitalized, moderate, severe, and deceased individuals across days from symptom onset. Points represent a unique individual-timepoint and color represents the clinical severity status of that given individual-timepoint as labeled. Moderate and severe classification represents only individual-timepoints of patients who survived. A cubic spline fitted to the mean with a lambda of 0.05 is shown for each severity group. Shading of the lines represent the bootstrap confidence of fit. **h**, Least squares Mean analysis comparing mean saliva viral load amongst spectrum of disease over days from symptom onset using Restricted Maximum Likelihood (REML) to account for repeated measures for the same individuals. Significance was adjusted for multiple comparisons using the Tukey method. **i**, Linear regression analysis, assessing correlations of saliva viral load between alive and deceased groups over days from symptom onset. Pearson’s correlation coefficients are shown for each group; shading represents the 95% confidence interval for the regression line. **j, k, l**, Comparison of nasopharyngeal viral load between spectrums of disease severity as described for saliva viral load in **g, h, and i** respectively. Dotted lines VLP=positivity threshold for viral load is 3.75 Log (Genomic Equivalents [GE]/ml)); VLD=viral load limit of detection is 3.22 Log (GE/ml).

To gain a better sense of how this played out throughout the course of disease we tracked saliva viral load levels over days from symptom onset. This revealed distinct viral load kinetics across all severity groups, with those with more severe disease initiating with or maintaining higher viral load levels throughout the course of disease (Figure 2g). Measurement of overall saliva viral load levels over time revealed that there were significant differences across all severity groups with viral load levels increasing significantly from non-hospitalized individuals, to moderate, severe, and finally deceased individuals (Figure 2h). Surviving patients showed a significant steady decline of viral load over time while those with fatal disease failed to control saliva viral load (Figure 2i). While this observation also held true for nasopharyngeal viral load (Figure 2l), all other comparisons did not significantly differ across severity groups when measured by nasopharyngeal viral load (Figure 2d-f, j, k).

We therefore performed a Receiver Operating Curve (ROC) analysis for both nasopharyngeal and saliva viral load which revealed that while nasopharyngeal viral load could not, saliva viral load could significantly predict both disease severity (AUC=0.71; p=0.0012) and mortality (AUC=0.83; p<0.0001) in hospitalized patients based on the first collection sample from the patient, even as this varied by several days past the date of admission and showed statistically better predictive ability than nasopharyngeal viral load and age (Extended Figure 2a and Extended Table 2). Incorporation of days from symptom onset significantly improved the predictive value of saliva viral load for both disease severity (AUC=0.79, p<0.0001) and mortality (AUC=0.90, p<.0001) (Extended Figure 2b, c) which was again significantly better than the nasopharyngeal viral load model (Extended Figure 2d) in predicting mortality. Using ordinal logistic regression, we tested if either saliva or nasopharyngeal viral load combined with days from symptom onset could predict the full spectrum of severity. Saliva viral load with days from symptom onset alone showed a strong predictive ability to distinguish the full spectrum of severity with first sample collection distinguishing between moderate disease (AUC=0.96) severe (AUC=0.89), and fatal (AUC=0.91) COVID -19 (Extended Figure 2b). These results highlight the distinctive feature of saliva viral load over nasopharyngeal viral load to correlate with a spectrum of disease severity and mortality throughout the course of disease and further suggest an important role for early viral load in determining outcomes of severity.

### Saliva viral load correlates with key immunological factors in COVID-19

To gain a better understanding of the association of viral load with disease severity and mortality, we investigated the overall relationship between saliva and nasopharyngeal viral load to several immunological factors measured in our cohort which included cytokines, chemokines, platelets, and antibody levels across all patient timepoints. Observation of these correlations revealed that, with notable exceptions, saliva load and nasopharyngeal load shared many positive and some negative correlates (Figure 3a).

**Fig. 3.**
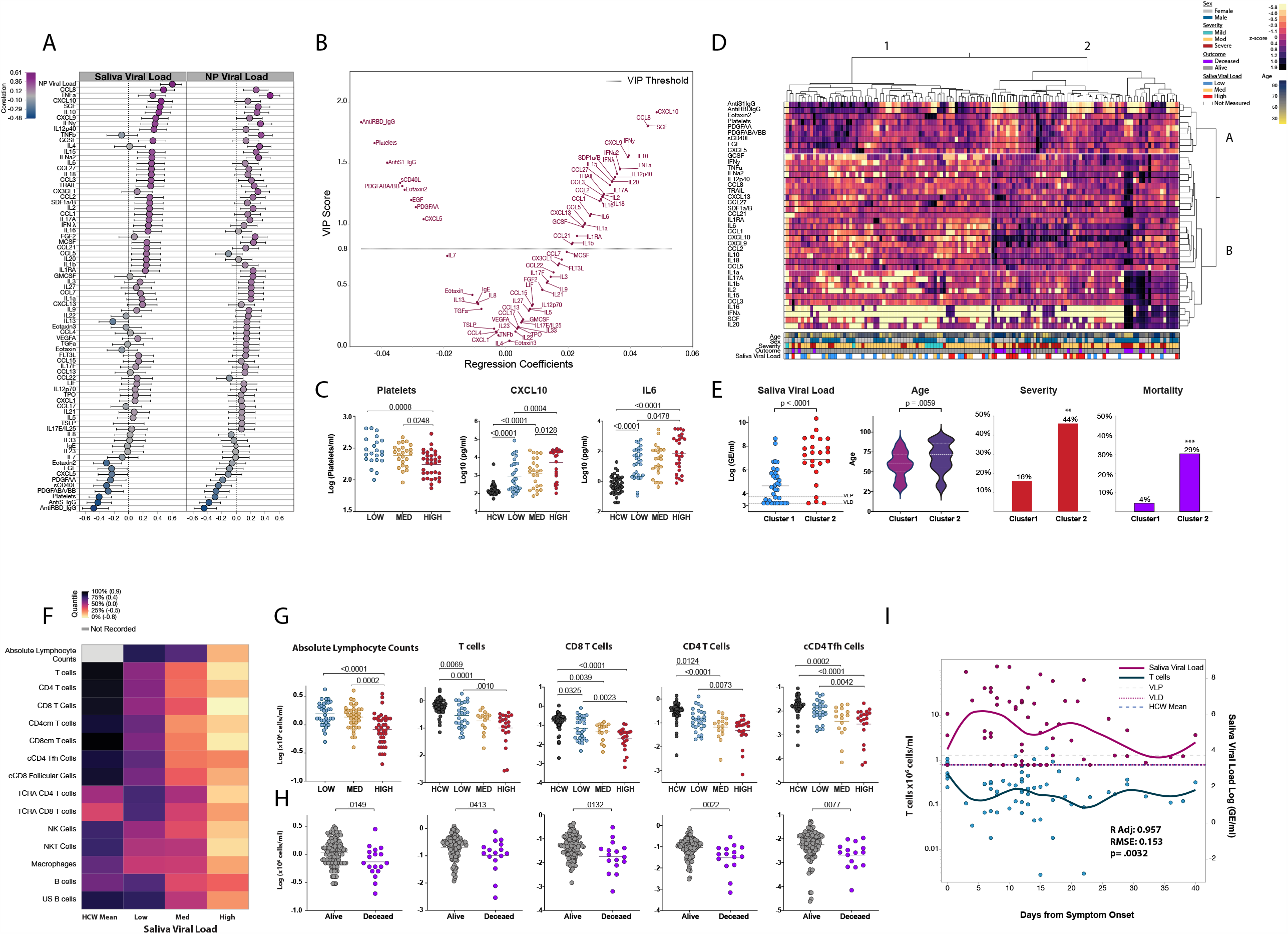
Saliva viral load correlates with key immunological markers of severity and mortality in COVID-19. **a-i, a**, Comparison of saliva and nasopharyngeal viral load to 75 immunological factors using Pearson correlation. Values are ordered starting at the top by maximum positive correlation value. Correlation scale is represented on the x-axis. Whiskers represent the 95% confidence interval of the correlation. **b**, Variable Importance Plot (VIP) vs coefficients plot generated from non-linear iterative partial least squares (NIPALS) analysis, with K-Fold cross validation (K=5). The plot represents component 1 which accounted for approximately 16.59% of the x variability and approximately 52% of the y (see Extended Figure 3a). The VIP threshold cutoff was 0.8. VIP score is denoted on the y-axis with regression coefficients on the x-axis. A total of 39 factors met the threshold criteria for importance; 30 of which were positive predictors of higher saliva viral load, and 9 of which were predictors of lower saliva viral load. **c**, One-way ANOVA comparing the mean difference of platelets, CXCL10, and IL-6, between healthcare workers (HCW) and/or individual timepoints in the low, medium, and high saliva viral load groups. P-values were adjusted using Tukey method. Additional comparisons are available in Extended Figure 4. **d**, heatmap depicts hierarchical clustering comparing enrichment of the 39 VIP factors identified in B measured at distinct timepoints in COVID-19 patients. Measurements were normalized across all patients. Clustering was Ward based (Cluster 1, n=68, cluster 2, n=62). **e**, Comparisons between clusters 1 and 2 generated from D. Comparison of saliva viral load and age, by 2-sided t-test; significance for frequency of severity and mortality was done via 2-tailed Fisher’s exact test and was p=0.0033, and p=0.0002 respectively. **f**, Heatmap depicting mean values of lymphocyte subset counts between healthcare workers, and low medium and high saliva viral load groups. Lymphocyte subsets are labeled or abbreviated on the left. Abbreviations: CD4cm T cells=CD4 central memory T cells; CD8cm T cells=CD8 central memory T cells; TCRA CD4 T cells=acutely activated effector CD4 T cells (HLADR+, CD38+, CD8 4+); TCRA CD8 T cells=acutely activated effector CD8 T cells (HLADR+, CD38+, CD8 +); US B cells=non-class switched memory B cells. **g**, One-way ANOVA comparing the mean difference of lymphocytes, and T cell subsets between uninfected healthcare workers (HCW) and/or individual-timepoints in the low, medium, and high saliva viral load groups. P-values were adjusted using Tukey method. Additional comparisons are available in Extended Figure 5a. **h**, Comparison of cell populations in G between alive and deceased individuals. P-values are adjusted for multiple comparisons using the Holm-Sidak method. Additional comparisons are available in Extended Figure 5c. **i**, Restricted Maximum Likelihood (REML) Least squares regression was performed correlating saliva viral load with T cells numbers in across time R adjusted=0.957, RMSE=0.153, p=0.0032) Additional comparisons are shown in Table 4. Dual-y axis graph tracking T cell counts (x10^6^ cells/ml) on the left and saliva viral load (Log GE/ml) on the right over days from symptom onset for all patients. Points represent a unique timepoint measurement for either t cell counts or saliva viral load as indicated on the graph. All points depicted for either T cell counts, or saliva viral load have a corresponding measurement of saliva viral load, or T cell counts respectively. Saliva viral load and T cell count curves are color coded as indicated. Curves represent the cubic spline fitted to the mean of the points with a lambda of 0.05. Dotted lines VLP=positivity threshold for viral load is 3.75 Log (Genomic Equivalents [GE]/ml)); VLD=viral load limit of detection is 3.22 Log (GE/ml). HCW mean=Mean T cell count for healthcare workers is also shown for reference.

Viral load obtained from either collection method positively correlated with many important markers of inflammation previously seen with nasopharyngeal viral load^8,33^, including IFN-α, and IFN-γ, inflammasome cytokines (IL-1β, IL-1ra), cytokines (TNFα, IL-6, IL-10), as well as with CXCL10 and many other chemokines responsible for the recruitment of monocytes, T cells, and other leukocytes, but showed stronger or significant associations with saliva load than with nasopharyngeal viral load (Figure 3a). Saliva viral load also showed statistically significant positive correlations with IL-18, and IFNλ, which were previously shown to be associated with disease severity and mortality^8^ but not previously identified to be directly correlated with viral load (Figure 3a). Both nasopharyngeal and saliva viral load showed a strong negative correlation with anti-S and anti-RBD IgG, platelets, as well as certain growth and repair factors.

Given the strong associations of saliva viral load over nasopharyngeal viral load with many of the immunological factors associated with increased disease severity and mortality, we sought to identify the most influential predictors of saliva viral load by running a non-linear iterative partial least squares (NIPALS) analysis of all factors and then assessing for variable importance. Variable Importance Plot (VIP) analysis revealed a total of 39 factors above the VIP threshold cutoff, 30 of which were positively associated with saliva viral load and 9 of which were negatively associated (Figure 3b). CXCL10, a chemokine that attracts activated T cells expressing CXCR3, was identified as the most important positive correlate of higher saliva viral load. In close second through fourth were additional chemokines and mitogens including CCL8, CXCL9, and SCF. Other positive VIP correlates once again included inflammatory and inflammasome cytokines (TNFα, IL-18, IL-1ra, IL-1α and IL-1β), cytokines (IL-10, IL-6, IL-15), chemokines (CXCL9, CCL3, CCL2), types I-III interferons, as well as additional Th1 and Th17 cytokines such as IL-2, IL-12p40, and IL-17a. CXCL13, a chemokine for Tfh and germinal center B cells expressing CXCR5, was also found to be a positive predictor of higher saliva viral loads.

The most important negative correlates of saliva viral load were found to be anti-RBD IgG, platelets, and anti-S IgG (Figure 3b), suggesting an important role for a humoral response against viral replication. Platelets might correlate negatively because of their depletion in severe disease that are seen with high viral load (Figure 3c, Extended Figure 5c). Interestingly, after these antibodies, the negative VIP correlates of saliva viral load—Eotaxin 2, PDGF ABA/BB, PDGFAA, EGF, sCD40L, and CXCL5—were factors derived from platelets or important in antibody class switch and found to be enriched in the plasma of patients who recovered from COVID-19 ^8^.

Comparing these VIP correlates between low medium and high viral load, we found that they followed either progressive decline or increase, depending on whether they were predictors of low or high viral load, respectively (Figure 3c, and Extended Figure 3b and 4). To better visualize how each of these positive and negative parameters of saliva viral load behaved within our cohort, a heatmap was constructed using these 39 VIP predictors of saliva viral load. Hierarchical clustering revealed two distinct patient clusters (cluster 1, and 2; Figure 3d) and two distinct clusters of cellular factors (A, B; Figure 3d). Patients in cluster 1 showed a strong enrichment of factors in cluster A, which were all of the VIP correlates associated with low saliva viral load, while patients in cluster 2 showed strong enrichment of factors in cluster B, which were positive predictors of high saliva viral load. Indeed, patients in cluster 2 showed significantly higher saliva viral loads, were significantly older, and showed significantly higher rates of severity and mortality (Figure 3e). These findings suggest that the pro-inflammatory signature seen in severe COVID-19 may be driven by higher viral loads. Further, they reveal an immunological blueprint associated with lower viral loads, decreased severity, and mortality.

### Saliva viral load is strongly associated with the progressive depletion of lymphocytes

One of the hallmarks of COVID-19 is T lymphopenia. We therefore investigated the association of viral load to the amount of T cells and other cell subsets found in patient PBMC samples. As previously described for other types of viral load^33^, saliva viral load was significantly negatively associated with absolute lymphocyte counts obtained from patient clinical data, as well as total T cells counts of CD4 and CD8 T cells (Extended Figure 3c). Additionally, saliva viral load was negatively associated with many effector T cell subsets, NK, and NKT cells, macrophages, and some B cell subsets (Extended Figure 3c). In contrast, nasopharyngeal viral load also showed some but fewer significant negative correlation to some of these lymphocyte subsets (Extended Figure 3c).

Comparison of lymphocyte subset counts to uninfected healthcare workers and individuals with low, medium, and high saliva viral load levels showed that individuals with high saliva viral load had significantly lower total T cell counts compared to uninfected healthcare workers and those in the low saliva viral load group (Figure 3f, g). These associations held even after accounting for age-related T cell decline (Extended Table 3). Similar observations were seen for CD4 and CD8 T cells and effector subsets including lower counts for CD4 Tfh and follicular CD8 T cells as well as lower counts for HLADR+/ CD38+ activated CD4 and CD8 T cells with higher saliva viral loads (Figure 3g, and Extended Figure 5a). Furthermore, patients in the high saliva viral load group exhibited significant reduction of NK cell, NKT cell counts over uninfected healthcare workers (Extended Fig. 5A). Similar effects were seen with nasopharyngeal viral load levels but not for all subsets including T cells when ranked by viral load (Extended Figure 5b).

Given the important relationship between days from symptom onset to viral load, we evaluated the relationship of saliva viral load to T cell counts over the course of disease. Saliva viral load, and days from symptom onset showed a significant contribution to the changes in T cell counts over time (Adj R=0.957, p=0.0032) (Figure 3i and Extended Table 3). These strong negative associations between saliva viral load over days from symptom onset extended to CD4 and CD8 T cells and effector subsets including circulating CD4 Tfh cells (Adj R=0.961, p=0.00045) (Extended Table 3). Nasopharyngeal viral load also showed some of these associations, though to a limited set of cells (Extended Table 3)

Generating highly specific anti-RBD IgG, the most important negative correlate of saliva viral load that we measured, requires the coordinated efforts of germinal center B cells as well as Tfh cells for antibody affinity maturation. Given the strong negative association that saliva viral load has on the numbers of circulating lymphocytes and lymphocyte subsets including CD4 Tfh cells, an assessment of the lymphocyte repertoire in deceased patients was conducted. This analysis revealed that deceased patients indeed exhibited significant depletion of circulating lymphocytes and T cell subsets including CD4 Tfh cells (Figure 3h and Extended Figure 5c).

### Early escape from high saliva viral load is associated with better antibody production and recovery

Anti-RBD IgG and anti-S IgG levels were among the most important negative correlates of low saliva viral load. Comparison of antibody levels between uninfected healthcare workers and those in the high medium and low saliva viral load levels demonstrated that patients with medium and low saliva viral load had significantly higher anti-RBD and anti-S IgG than those in the high viral load group (Figure 4a, d). To gain a better understanding of the temporal association of antibody production to viral load, antibody levels were compared amongst patients in the high saliva viral load level to those with lower saliva viral load levels over time. Patients who stayed in the high saliva viral load level showed an initial delay in the production of antibody compared to those in the lower saliva viral load levels with a significant difference between days 10-20 from symptom onset, before eventually catching up to those who were in, or had managed to enter into the lower viral load levels (Figure 4b, e). Notably, this statistically significant difference in antibody production between days 10-20 from symptom onset was also seen in deceased individuals (Figure 4 c, f), which coincided with increased depletion of cCD4 Tfh cells during this time period (Extended Figure 4a).

**Fig. 4.**
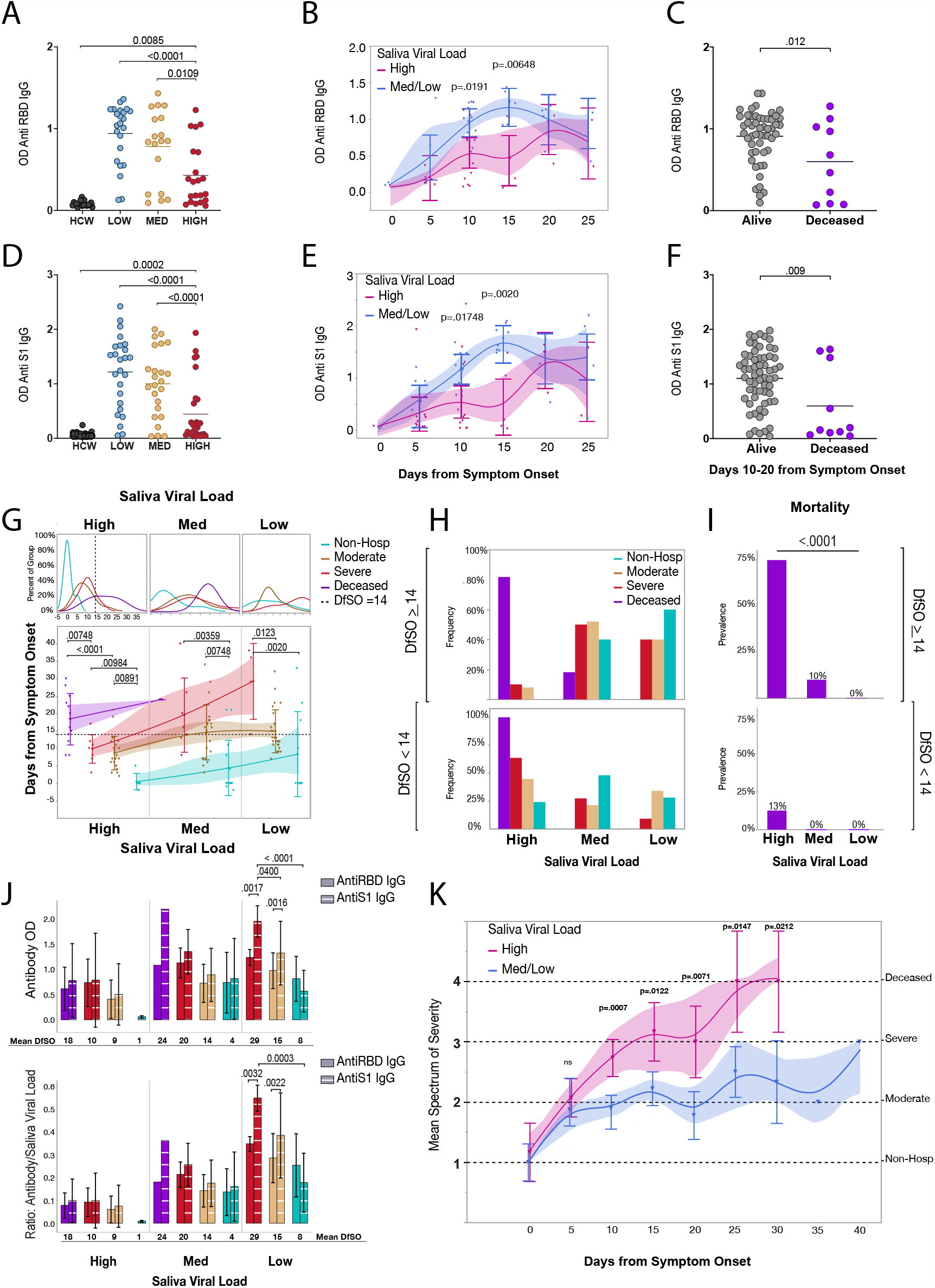
Early escape from high saliva viral load is associated with better antibody production and recovery. **a-k, a**, Comparison of mean anti-RBD IgG levels between low, medium, and high saliva viral Load and uninfected healthcare workers. Comparisons were done using a one-way ANOVA p-values were adjusted using Tukey method for multiple comparisons. **b**, Least squares means analysis comparing Anti-RBD IgG levels amongst patients with high vs medium and low saliva viral load over days from symptom onset. Timepoints represent 5-day bins. P values are adjusted for multiple comparisons using Sidak correction. Whiskers and shading represent the 95% confidence interval of the mean. **c**, Comparison of the mean Anti-RBD IgG levels amongst alive and deceased individuals between days 10-20 from symptom onset using a 2-sided t-test and. P-vaules are adjusted for multiple comparisons with Sidak correction. **d, e, f**, Comparison of Anti-S1 IgG levels as in in **a, b, c** respectively. **g**, Comparison of the mean duration of days from symptom onset in the high, medium and low saliva viral load categories amongst spectrum of disease severity. <u>Top Panel</u> shows the Kernel projected percent distribution of time spent for individuals within severity subgroups in the high, medium, or low saliva viral load categories. <u>Bottom panel</u> compares the mean time (y -axis) of each severity subgroup within each saliva viral load category. Points represent a patient time point. Severity is color-coded as indicated on the graph. Lines connect the means within each severity subgroup in each viral load category. Whiskers represent the standard deviation of the mean and shading represents the 95% bootstrap confidence of fit. Comparisons within each viral load category were done by one-way ANOVA and significance was adjusted for multiple comparisons across all categories using the Holm-Sidak method. The dotted line in the top and bottom panels labeled as high viral load Escape Threshold in the legend (Day 14 from symptom onset) represents the rounded average of one standard deviation away from the mean for the moderate and severe groups of patients in the high saliva viral load category who survived. <u>**h**</u>, Panels are subdivided by the high viral load Escape threshold (Day 14 from symptom onset) as indicated in the Figure. Panels represent the frequency of individuals within each severity group that fall into the high, medium, and low saliva viral load categories before day 14 from symptom onset (bottom) and at or after day 14 from symptom onset (top). i, Subpanels show the prevalence of individuals who go on to die within the high, medium, and low saliva viral load categories before day 14 (bottom) and on or after day 14 (top). Chi-squared tests were used to measure significance. j, Panels show mean levels of Anti S1 IgG and Anti RBD IgG (top) and their ratios over saliva viral load (bottom), amongst severity groups in the high medium and low saliva viral load levels. X-axis denotes saliva viral load levels as well as mean days from symptom onset (DfSO) of each severity group within each viral load level as determined in **g**. Comparisons of means amongst severity groups is made in the low saliva viral load level via least squares mean analysis using REML to compare Anti S1 IgG and Anti RBD levels between the groups and within groups. P values are adjusted for multiple comparisons using Sidak correction. There was a significant positive interaction between disease severity and higher proportions of Anti S1 IgG over Anti RBD IgG (p=0.0033). **k**, Least squares mean analysis comparing disease trajectory between high and medium/low viral loads over days from symptom onset. Disease severity was ranked on an increasing scale from 1-4 with 1 being illness not requiring hospitalization, 2 hospitalization with moderate disease, 3 severe disease, and 4 death. Timepoints represent 5-day bins. Points represent the mean score of disease severity for all patients in each bin that fell within the high vs medium/low saliva viral load categories. Whiskers represent the 95% confidence interval and shading represents the bootstrap confidence of fit. There was a significant positive interaction between viral load level and days from symptom onset (p=0.0187).

To better understand how antibody levels correlated with saliva viral load kinetics and disease severity in our cohort, the mean duration of time spent in the high, medium, and low saliva viral load brackets was compared amongst the spectrum of disease severity along with antibody levels and the overall ratio of antibody to saliva viral load levels. Interestingly, deceased patients exhibited significantly longer time spent in the high saliva viral load bracket than severe, moderate, or non-hospitalized individuals, while non-hospitalized individuals spent a significantly shorter time (less than 5 days) in the high viral load bracket than hospitalized patients (Figure 4g). While antibody levels did not vary significantly across the cohort in the high viral load bracket, the overall ratio of antibody to saliva viral load was uniformly low (Figure 4h). Notably the vast majority of patients who survived exited the high saliva viral load bracket by day 14 from symptom onset, which accounted for one standard deviation from the mean for both moderate and deceased individuals in the high saliva viral load group and fell within the significant window of antibody production described in Figure 4b. Furthermore, with the exception of two individuals in the medium viral load level, deceased patients failed to exit the high saliva viral load bracket (Figure 4g-j). Observation of individual trajectories for saliva viral load and antibody production, showed that saliva viral load decreased as overall antibody to viral load ratio increased within an individual (Extended Figure 6c). Notably, antiviral antibody to saliva viral load ratios in the medium and low viral load brackets showed a steady increase amongst the severity groups, with higher antibody and antibody to viral load ratios seen with increasing disease severity and lower viral load respectively (Fig 4j), as well as a growing separation between anti-S1 IgG and anti-RBD IgG that increased with disease severity and the mean days from symptom onset (Figure 4g, h). Indeed, patients with severe disease who successfully made it into the low viral load group did so at significantly more days from symptom onset than moderate or non-hospitalized individuals. In addition, they displayed significantly higher anti-S1 IgG levels than the lower severity groups (Figure 4h). Furthermore, higher proportional levels of anti-S1 IgG over anti-RBD IgG in the low saliva viral load group showed a significant positive interaction with increased disease severity, with Non-Hospitalized individuals showing a non-significant difference between their anti-RBD IgG and anti-S1 IgG levels, and those with moderate and severe disease showing significant and increasing differences in their anti-S1 IgG levels compared to anti-RBD IgG levels (Figure 4j). Of note, these associations were not seen with nasopharyngeal viral load (Extended Figure 6 d-g).

Together these data suggest two important features of viral load and antibody in the determination of disease severity and mortality in COVID-19. First, patients with higher initial viral load levels required higher concentrations of antibody, and higher antibody-to-viral-load ratios to escape the high and medium saliva viral load bracket and effectively overcome their viral burden. Second, while anti-RBD IgG was significantly more effective at bringing down saliva viral load (Extended Figure 6b), increased disease severity required broader anti-S1 IgG to overcome their viral burden (Figure 4j, which may further delay their overall rate of recovery. Given that higher viral loads are associated with depletion of cCD4 Tfh cells, these results suggest that lethal COVID-19 disease may be a feature of high early stage saliva viral load, reduction in Tfh and antibody responses, leading to the failure to clear the virus, and triggering a vicious circle. Indeed, maintaining high viral load for longer showed significantly divergent trajectories toward increasing severity of illness and death (Fig. 4k).

## Discussion

Our study, consistent with others who have shown differences in viral load associations amongst different collection sites^19,33^, revealed that nasopharyngeal and saliva viral load, are not equivalent measures of disease processes for COVID-19. Correlation between these two sites showed an R=0.61 (p<0.0001) and saliva and nasopharyngeal viral load shared a related but an overall distinct cytokine signature. Saliva viral load was significantly higher with increasing disease severity and demonstrated distinct viral load kinetics across the severity spectrum from non-hospitalized to deceased individuals. Saliva viral load could significantly predict disease severity and mortality over age or nasopharyngeal viral load with the first sample collection, and, importantly, was enhanced with the inclusion of days from symptom onset. Nasopharyngeal viral load could not reliably distinguish severity or predict mortality in our study with the first sample collection—which varied from the date of admission. This distinction is important in consideration of prognostic tools that may be used clinically to monitor disease progression after the date of admission. These findings suggest that saliva viral load is reflective of a related but distinct replicative process in vivo than what is seen with nasopharyngeal viral load. Though nasopharyngeal and oral epithelia as well as epithelia surrounding salivary glands show ACE2 expression^36,37^, a replicative capacity for SARS-CoV-1^38^, and one study found pronounced ACE2 expression in minor salivary glands of humans infected with SARS-CoV-2^39^, the collection of virus in saliva may also be reflective of viral replication in key vital organs. Saliva viral load may reflect viral invasion and replication in the lower respiratory tract that has been propelled into the oral cavity through mucociliary clearance that subsequently intermixes with saliva^40^. Nevertheless, the significant and temporal association of saliva viral load with spectra of disease severity suggests that early high viral loads associated with saliva are the immunological trigger for disease severity and those who sustain high viral loads longer suffer increased disease severity or mortality.

The association of older age, heart failure, cancer, and certain forms of immunosuppression with higher viral loads suggests a possible interplay between these factors^1-4,27^. Our study evaluating saliva not only confirms the association of higher viral load with these and other known COVID-19 risk factors including hypertension, and chronic lung disease, but demonstrates that these factors have an additive effect in overall viral load levels, which may contribute to the increased risk of severity in individuals with multiple risk factors^4^. Our study also found that saliva viral load was significantly higher in males, a known risk factor in COVID-19, associated with a distinct immune response than what is seen in females^3,41-43^. Therefore COVID-19 risk factors may be reflective of a baseline capacity of the host to support viral replication, predisposing those with multiple risk factors to higher viral loads and worse outcomes. The high early viral replication in a given host is likely a reflection of failed innate immune responses including interferon-mediated antiviral defense. Indeed, impaired innate antiviral defense is seen with the COVID-19 risk factors including advanced age^44^, metabolic syndrome^45^, cancer^46^,and in immunocompromised status. In addition, recent work has also shown increased expression of ACE2 in the lungs of individuals with COVID-19 risk factors^47^, which may further predispose those with pre-existing conditions to higher viral replication.

Thirty-nine VIP factors were significantly associated with saliva viral load, including 30 factors associated with higher saliva viral load, which included Type I-III IFNs, inflammasome factors, cytokines and chemokines such as CXCL13; and 9 associated with lower viral loads. These two signatures associated with high and low saliva viral load clustered separately from one another in our cohort and those who had enrichment of the lower viral load signature, which included six factors (Eotaxin 2, PDGF ABA/BB, PDGFAA, EGF, sCD40L, and CXCL5) previously associated with recovery in our cohort ^8^, showed significantly lower rates of disease severity and mortality. Many of these lower viral load factors are released by platelets which also showed a significant predictive ability for lower saliva viral loads. This phenomenon may be related to the thrombocytopenia and coagulopathies seen in patients with severe disease that is reflected through saliva viral load and further integrates saliva viral load as an important measure of the disease phenotypes seen in COVID-19. These results provide an immunological blueprint that reflects better protection against viral replication; a path that may be inefficiently achieved in those who start with higher viral loads and produce higher inflammatory signatures.

Patients within the higher initial viral load bracket required higher concentrations of antibody and higher antibody-to-viral-load ratios to successfully escape the high viral load bracket. While anti-RBD IgG is more effective at bringing down viral load, increased disease severity was associated with higher reliance on less specific anti-S1 IgG to overcome their viral load burden, which may further delay their overall rate of recovery. Indeed, patients with severe disease took significantly longer days from symptom onset than those with moderate disease or non-hospitalized individuals to achieve low saliva viral loads. Given that higher saliva viral loads are associated with increased depletion of CD4 cTfh cells, these results suggested that this increased reliance on anti-S1 IgG may be driven from the lack of proper B cell help, as a result of lymphocyte depletion among this cohort.

In this same vein, patients with fatal COVID-19 showed significantly lower lymphocytes and lymphocyte subsets including CD4 cTfh cells than patients who survived. Like individuals with high saliva viral load, deceased patients exhibited significantly lower anti-RBD and anti-S1 IgG in days 10-20 from symptom onset, which corresponded with a significantly more pronounced depletion of cTfh cells in that time frame. This significant delay in antibody production may indeed have been the final determinant of their overall fate, for not only would they have to catch up in the production of antibody, but they would have to produce much more antibody than those with lower viral loads to effectively overcome their viral load burden and exit the high saliva viral load bracket. Concordantly, while most patients who survived successfully escaped the high saliva viral load bracket by day 14 from symptom onset, deceased individuals failed to do so showing low overall antibody to viral load ratios, and those that did manage to escape at later days showed signs of requiring much higher antibody levels to do so with a stronger reliance on anti S1 IgG. Hence, lethal COVID-19 disease may be a feature of high early stage saliva viral load, reduction in Tfh and antibody responses, leading to the failure to clear the virus, and triggering a vicious circle. Indeed, maintaining high viral load for longer or transitioning out of high saliva viral load showed significantly distinct trajectories toward recovery or death.

These findings may serve as a window to apply and monitor interventions dedicated to reducing viral load early before severe depletion of effector cell populations in days 10-20 from symptom onset which may contribute to the significantly lower levels of antibody seen during this time in the deceased. While some drugs and monoclonal antibodies designed to reduce overall viral load, have shown little-to-mixed efficacy in trials, this may be related to the inability of these drugs to successfully bring down viral burden at the early stage of disease^48,49 50,51^. It is also important to note that those who have higher viral loads required a higher antibody-to-virus ratio to successfully bring down viral load and anti-RBD IgG was better at accomplishing this. These findings are important when considering the incorporation of convalescent serum in individuals which may harbor varying levels of specific antibody and may successfully or unsuccessfully bring down viral loads to an appropriate ratio based on initial viral load levels. Serial monitoring of saliva viral load is important in these circumstances and others that target viral load to ensure efficacy.

In summary our work revealed both the important role that saliva viral load has as a clinical measure of disease severity and mortality in COVID-19 as well the importance of viral load as an integral component of the spectrums of disease severity and clinical pathology that should be monitored early. We identified novel factors associated with lower and higher viral loads that may serve as a basis for treatment, and highlight the importance of antibody responses against infection. Early interventions directed against viral load may be most beneficial if administered early before days 10-20 from symptom onset to prevent increased viral load associated depletion of effector subsets and corresponding decrease in antibody production, allowing for a more coordinated immune response against the virus.

## Methods

### Study Cohort and data collection

#### Patients

One-hundred and fifty-four SARS-CoV-2 RT-q-PCR positive eligible^34,41^ adult (≥ 18 years old) patients admitted to Yale New Haven hospital between 18 March 2020 to 10 June 2020 were included in this study. Patients were confirmed to be positive by at least one SARS-CoV-2 PCR-positive nasopharyngeal swab, collected during or near the time of admission. Patients were followed longitudinally throughout the duration of their hospital stay. Disease severity was determined based on the patient’s overall requirement of medical intervention at each timepoint of data collection, and were categorized into the following severity groups: (i) “non-hospitalized”, consisting of patients who were admitted for elective care and were discovered to be positive via nasopharyngeal swab RT-qPCR screening but who did not require medical intervention for their COVID-19 infection throughout their stay; (ii) moderate disease, consisting of patients admitted to the hospital for their COVID-19 illness and requiring only non-intensive level of medical care, up to the administration of non-invasive supplementary oxygen (>3 L/min to maintain SpO_2_ >92%, or >2 L/min to maintain SpO_2_ >92% with a high-sensitivity C-reactive protein [CRP] >70), plus treatment with tocilizumab; (iii) severe, which included patients who were admitted to the intensive care unit (ICU) and required intensive level of medical care including requiring >6 L/min of supplementary oxygen to maintain SpO_2_ >92%, invasive mechanical ventilation or extracorporeal membrane oxygenation (ECMO) in addition to glucocorticoid or vasopressor administration; (IV) deceased patients, consisting of patients admitted for COVID-19 and died during their hospital stay. Patient demographic information and COVID-19 health risk factors including information presented in Extended Table 1 were obtained from a systematic and retrospective review of electronic medical records (EMRs). Days from symptom onset for each patient was determined via a standardized interview and collected on a standardized survey administered to the patient or via review of electronic medical records if an interview was not possible due to patient’s health or mental status. All clinical data were obtained using EPIC EHR and REDCap 9.3.6 software. Investigators were blinded to patient information and health status at the time of sample processing and generation of raw data from blood and plasma samples. Blood specimen collection and specimen processing were performed by independent teams. Cytokines and FACS analyses were blinded. Clinical data including absolute lymphocyte counts and platelets were obtained from the patient’s EMR and were matched to the date of or a maximum of 48 hours from saliva or nasopharyngeal swab sample collection. Patients clinical information and clinical severity status was only revealed after the generation of raw data. Of note, our cohort consists of individuals who received Tocilizumab, which can increase the levels of IL-6 as previously described^8^. **Healthcare workers:** One hundred and nine uninfected healthcare workers were used in this study as a control group. Age and sex were obtained from standardized survey data. Healthcare workers were serially monitored for infection using RT-qPCR screening from either nasopharyngeal or saliva samples. Eighteen healthcare workers who became RT-q-PCR positive for SARS-CoV-2 but did not require hospitalization for symptoms were included in this study as part of the non-hospitalized group. Days from symptom onset were determined from a standardized survey provided to healthcare workers throughout enrollment in the study, or by interview.

#### Plasma and PBMC isolation

Patient and healthcare worker plasma and PBMC isolation was obtained as previously described^8,41^. Briefly, same-day blood collection and processing was done from whole blood samples collected in sodium heparin-coated vacutainers that were gently agitated until the samples were processed. To isolate serum from blood samples, vacutainers were centrifuged at 400g for 10 min at room temperature (RT) with no brake. The resulting undiluted serum was aliquoted and stored at −80 °C for later analysis. For PBMC isolation, resulting concentrated blood after serum isolation was processed at room temperature, first by diluting the sample in a 1:1 ratio with PBS. And then by density gradient centrifugation using 50 ml SepMate tubes (StemCell Technologies; #85460) in which Histopaque (Sigma-Aldrich, #10771-500ML) had been added to the appropriate volume according to the manufacturer’s instructions. Samples were then centrifuged for 10 mins at 1,200g with the brake on. Resulting PBMC layer was decanted into a new sterile 50ml conical tube and washed twice with PBS, treated with ACK lysis buffer for 2 min and washed again before counting and assessing cell viability using standard Trypan blue staining and an automated cell counter (Thermo-Fisher, #AMQAX1000). All samples were processed in a BSL2+ level facility.

#### Cytokine and chemokine measurements

Cytokine and chemokine data were obtained as previously described^8,41^. Stored patient sera (as described above) were shipped to Eve Technologies (Calgary, Alberta, Canada) on dry ice for the quantification of 71 cytokines and chemokines using the Human Cytokine Array/Chemokine Array 71-Plex Panel. Samples were measured at first-thaw.

#### Flow Cytometry

Flow Cytometry was performed as previously described^8,41^. Antibody clones and vendors used for flow cytometry staining were as follows: BB515 anti-hHLA-DR (G46-6) (1:400) (BD Biosciences), BV785 anti-hCD16 (3G8) (1:100) (BioLegend), PE-Cy7 anti-hCD14 (HCD14) (1:300) (BioLegend), BV605 anti-hCD3 (UCHT1) (1:300) (BioLegend), BV711 anti-hCD19 (SJ25C1) (1:300) (BD Biosciences), AlexaFluor647 anti-hCD1c (L161) (1:150) (BioLegend), biotin anti-hCD141 (M80) (1:150) (BioLegend), PE-Dazzle594 anti-hCD56 (HCD56) (1:300) (BioLegend), PE anti-hCD304 (12C2) (1:300) (BioLegend), APCFire750 anti-hCD11b (ICRF44) (1:100) (BioLegend), PerCP/Cy5.5 anti-hCD66b (G10F5) (1:200) (BD Biosciences), BV785 anti-hCD4 (SK3) (1:200) (BioLegend), APCFire750 or PE-Cy7 or BV711 anti-hCD8 (SK1) (1:200) (BioLegend), BV421 anti-hCCR7 (G043H7) (1:50) (BioLegend), AlexaFluor 700 anti-hCD45RA (HI100) (1:200) (BD Biosciences), PE anti-hPD1 (EH12.2H7) (1:200) (BioLegend), APC anti-hTIM3 (F38-2E2) (1:50) (BioLegend), BV711 anti-hCD38 (HIT2) (1:200) (BioLegend), BB700 anti-hCXCR5 (RF8B2) (1:50) (BD Biosciences), PE-Cy7 anti-hCD127 (HIL-7R-M21) (1:50) (BioLegend), PE-CF594 anti-hCD25 (BC96) (1:200) (BD Biosciences), BV711 anti-hCD127 (HIL-7R-M21) (1:50) (BD Biosciences), BV421 anti-hIL17a (N49-653) (1:100) (BD Biosciences), AlexaFluor 700 anti-hTNFa (MAb11) (1:100) (BioLegend), PE or APC/Fire750 anti-hIFNy (4S.B3) (1:60) (BioLegend), FITC anti-hGranzymeB (GB11) (1:200) (BioLegend), AlexaFluor 647 anti-hIL-4 (8D4-8) (1:100) (BioLegend), BB700 anti-hCD183/CXCR3 (1C6/CXCR3) (1:100) (BD Biosciences), PE-Cy7 anti-hIL-6 (MQ2-13A5) (1:50) (BioLegend), PE anti-hIL-2 (5344.111) (1:50) (BD Biosciences), BV785 anti-hCD19 (SJ25C1) (1:300) (BioLegend), BV421 anti-hCD138 (MI15) (1:300) (BioLegend), AlexaFluor700 anti-hCD20 (2H7) (1:200) (BioLegend), AlexaFluor 647 anti-hCD27 (M-T271) (1:350) (BioLegend), PE/Dazzle594 anti-hIgD (IA6-2) (1:400) (BioLegend), PE-Cy7 anti-hCD86 (IT2.2) (1:100) (BioLegend), APC/Fire750 anti-hIgM (MHM-88) (1:250) (BioLegend), BV605 anti-hCD24 (ML5) (1:200) (BioLegend), BV421 anti-hCD10 (HI10a) (1:200) (BioLegend), BV421 anti-CDh15 (SSEA-1) (1:200) (BioLegend), AlexaFluor 700 Streptavidin (1:300) (ThermoFisher), BV605 Streptavidin (1:300) (BioLegend). Briefly, freshly isolated PBMCs were plated at 1–2 × 10^6^ cells per well in a 96-well U-bottom plate. Cells were resuspended in Live/Dead Fixable Aqua (ThermoFisher) for 20 min at 4 °C. After a wash, cells were treated with Human TruStan FcX (BioLegend) for 10 min at RT for blocking. Antibody cocktails were added directly to this mixture for 30 min at RT. For secondary stains, cells were washed and the supernatant was aspirated leaving a cell pellet. Then secondary cocktail of markers was added to the cell pellets for 30 min at 4 °C. Cells were resuspended in 100 μl 4% PFA for 30 min at 4 °C and then washed prior to data collection on an Attune NXT (ThermoFisher). FlowJo software version 10.6 (tree Star) was used for data analysis. The specific sets of markers used to identify each subset of cells are provided in Extended Figure 7.

#### Viral RNA measurements

Saliva samples and nasopharyngeal swab samples were obtained from patients and healthcare workers as previously described^34,52^. Collection of samples were attempted every few days for each patient throughout their stay. Samples were stored at room temperature and processed within 12 hours of sample collection. Nucleic acid was extracted from 300 μl of viral transport media from nasopharyngeal samples or extracted directly from 300 μl of saliva using a modified protocol for the the MagMAX Viral/Pathogen Nucleic Acid Isolation kit (ThermoFisher Scientific)^52^. 10 μl of Proteinase K was added as needed for some saliva samples which were then vortexed on high for 30 seconds to help further dissociate the sample for nucleic acid extraction. RNA was eluted using 75 μl of elution buffer. 5 μl of eluted RNA was used to SARS-Cov-2 detection using the US CDC real-time RT–qPCR primer/probe sets for 2019-nCoV_N1, 2019-nCoV_N2, and the human RNase P (RP) as an extraction control, under the protocol previously described^52^. Ct values obtained from the N1 primer-probe set were converted to viral genomic RNA copies per ml of sample input using a 10-fold dilution standard curve as described previously^52^. Values were Log_10_ transformed. The lower limit threshold for positive detection (viral load limit of positivity [VLP]) in our study was 5,610 virus RNA copies/ml of sample or approximately 3.75 Log_10_ (GE/ml). The viral load limit of detection (VLD) was 1,660 virus genomic RNA copies/ml or approximately 3.22 Log_10_ (GE/ml).

#### SARS-CoV-2 specific-antibody measurements

For the quantification of SARS-CoV-2 specific antibody ELISAs were performed as previously described^53^. Briefly, a final concentration of 0.5% and 0.5mg/ml of Triton X-100 and RNase A were added to serum samples respectively and incubated at room temperature (RT) for 30 minutes before use to neutralize any potential virus in serum. 96-well MaxiSorp plates (Thermo Scientific #442404) were coated with 50 μl/well of recombinant SARS Cov-2 S1 protein (ACROBiosystems #S1N-C52H3-100 μg) at a concentration of 2 μg/ml in PBS and were incubated overnight at 4 °C. After removal of the coating buffer, 200 μl of blocking solution (PBS with 0.1% Tween-20, 3% milk powder) was added to the plates and then subsequently incubated for 1h at RT. Serum was diluted in 1:50 dilution solution (PBS with 0.1% Tween-20. 1% milk powder) and 100 μl of diluted serum was added for two hours at RT. After three washes with PBS-T (PBS with 0.1% Tween-20) and 50 μl of HRP anti-Human IgG Antibody (GenScript #A00166, 1:5,000) or anti-Human IgM-Peroxidase Antibody (Sigma-Aldrich #A6907, 1:5,000) diluted in dilution solution added to each well. Plates were incubated for 1 h at RT, and then washed three times with PBS-T. Plates were developed with 100 μl of TMB Substrate Reagent Set (BD Biosciences #555214) and the reaction was stopped after 12 min by the addition of 2 N sulfuric acid. Plates were then read at a wavelength of 450 nm and 570 nm. Eighty pre-pandemic samples were assayed to establish the negative baselines and these values were statistically determined with a confidence level of 99%.

### Statistical and data analyses

Statistical and data analyses were performed using Jmp Pro 15.0.0 (SAS Institute), and GraphPad Prism 8.4.3. Parametric ANOVAs were performed as indicated in the figures for group analyses and p-values were adjusted for multiple comparisons as noted. Heatmaps were created using Jmp Pro 15.0.0. Measurements were normalized across all patients. Clustering was Ward based. Variable Importance Plot (VIP) vs coefficients plot was generated from non-linear iterative partial least squares (NIPALS) analysis, with K-Fold cross validation (K=5) using the Jmp Pro software.

## Data Availability

All the background information on HCWs, clinical information for patients, and raw data used in this study are included in Extended Table 4. Additionally, all of the raw .fcs files for the flow cytometry analysis are available

## Data availability

All the background information on HCWs, clinical information for patients, and raw data used in this study are included in <u>Extended Table 4.</u> Additionally, all of the raw .fcs files for the flow cytometry analysis are available at (not yet available).

## Acknowledgements

We thank M. Linehan for technical and logistical assistance, and P. Liu and W. Garcia-Beltran for discussions. This work was supported by the NIH Medical Scientist Training Program Training Grant T32GM136651, the Women’s Health Research at Yale Pilot Project Program, Fast Grant from Emergent Ventures at the Mercatus Center, Mathers Foundation, and the Ludwig Family Foundation, the Department of Internal Medicine at the Yale School of Medicine, Yale School of Public Health and the Beatrice Kleinberg Neuwirth Fund. IMPACT received support from the Yale COVID-19 Research Resource Fund. A.I. is an Investigator of the Howard Hughes Medical Institute. C.L. is a Pew Latin American Fellow. P.Y is supported by Gruber Foundation and the NSF. B.I. is supported by NIAID 2T32AI007517-16. C.B.F.V. is supported by NWO Rubicon 019.181EN.004.

## Author contributions

J.S., A.L.W. and A.I. conceived the study. C.L., J.K., J.S., J.E.O. and T.M collected and processed patient PBMC and plasma samples. P.W performed the flow cytometry and J.S. analysed the flow data. J.S., and B.I. collected epidemiological and clinical data. F.L. performed the SARS-CoV-2 specific antibody ELISAs. A.R. supervised the ELISAs. A.L.W., C.B.F., P.L., A.V., A.P., and M.T. performed samples processing, extractions, and RT-qPCR assays, under supervision of N.D.G. A.C.-M., M.C.M processed and stored patient specimens. J.Z. and A.V.W assisted in mild disease volunteer recruitment. M.C., J.B.F., C.D.C. and S.F. assisted hospitalized patients’ identification and enrolment. W.L.S. supervised clinical data management. J.S. and A.I. drafted the manuscript. All authors helped to edit the manuscript. A.I. secured funds and supervised the project.

## Yale IMPACT Research Team

Abeer Obaid^10^, Adam J. Moore^2^, Alice Lu-Culligan^1^, Allison Nelson^10^, Angela Nunez^10^, Anjelica Martin^1^, Anne E. Watkins^2^, Bertie Geng^10^, Chaney C. Kalinich^2^, Christina A. Harden^2^, Codruta Todeasa^10^, Daniel Kim^1^, David McDonald^10^, Denise Shepard^10^, Edward Courchaine^11^, Elizabeth B. White^2^, Eric Song^1^, Erin Silva^10^, Eriko Kudo^1^, Giuseppe DeIuliis^10^, Harold Rahming^10^, Hong-Jai Park^10^, Irene Matos^10^, Isabel M. Ott^2^, Jessica Nouws^10^, Jordan Valdez^10^, Joseph Lim^12^, Kadi-Ann Rose^10^, Kelly Anastasio^13^, Kristina Brower^2^, Laura Glick^10^, Lokesh Sharma^10^, Lorenzo Sewanan^10^, Lynda Knaggs^10^, Maksym Minasyan^10^, Maria Batsu^10^, Mary E. Petrone^2^, Maxine Kuang^2^, Maura Nakahata^10^, Melissa Campbell^6^, Melissa Linehan^1^, Michael H. Askenase^14^, Michael Simonov^10^, Mikhail Smolgovsky^10^, Nicole Sonnert^1^, Nida Naushad^10^, Pavithra Vijayakumar^10^, Rick Martinello^3^, Rupak Datta^3^, Ryan Handoko^10^, Santos Bermejo^10^, Sarah Prophet^15^, Sean Bickerton^11^, Sofia Velazquez^14^, Tyler Rice^1^, William Khoury-Hanold^1^, Xiaohua Peng^10^, Yexin Yang^1^, Yiyun Cao^1^, Yvette Strong^10^

^10^Yale School of Medicine, New Haven, CT, USA.

^11^Department of Biochemistry and of Molecular Biology, Yale University School of Medicine, New Haven, CT, USA.

^12^Yale Viral Hepatitis Program; Yale University School of Medicine, New Haven, CT, USA.

^13^Yale Center for Clinical Investigation, Yale University School of Medicine, New Haven, CT, USA.

^14^Department of Neurology, Yale University School of Medicine, New Haven, CT, USA.

^15^Department of Molecular, Cellular and Developmental Biology, Yale University School of Medicine, New Haven, CT, USA.

**Extended Data Fig. 1** |**Higher saliva viral load correlates with COVID-19 health risk factors. a-d, a**, Comparison of saliva viral load amongst individuals with and without certain COVID-19 health risk factor categories as denoted above each comparison and described in Extended Data Table 1. Significance for each t-test is adjusted for multiple comparisons using the Holm-Sidak method. **b**, Comparisons as in a, using nasopharyngeal viral load. **c, d**, frequency of disease severity or mortality respectively amongst individuals stratified by their cumulative totals of COVID-19 health risk factors from 0 to 4 or more health risk factors (4+) of those represented in **a** and **b**. Measure of significance for trend was determined using the Cochran Armitage Trend Test.

**Extended Data Fig. 2** | **Saliva viral load can significantly distinguish spectrums of disease severity and predict mortality a-d, a**, Receiver Operator Curve (ROC) of age, and first-recorded saliva and nasopharyngeal viral load values of hospitalized patients, as predictors of mortality. Area Under the Curve (AUC) was 0.83 for saliva viral load (p=0.0012), 0.63 (not significant[ns]) for age, and 0.58 (ns) for nasopharyngeal viral load. **b**, ROC models for mortality between saliva and nasopharyngeal viral loads combined with days from symptom onset are shown. AUC was 0.90 (p< 0.0001) for the saliva viral load model with days from symptom onset, and 0.67 for nasopharyngeal (p=0.06) viral load with days from symptom onset. AUC comparisons between models in a, and b were done using a chi-squared difference test and p-values for these comparisons are shown. **c, d**, ROC analysis demonstrating the predictive ability for saliva and nasopharyngeal viral loads respectively combined with days from symptom onset as predictors of the severity spectrum with only the first-obtained patient sample. Analysis was done using ordinal logistic regression. For saliva viral load with days from symptom onset (**c**) (p< .0001), the AUC was 0.91 for fatal disease, 0.89 for severe non-fatal disease, and 0.96 for moderate, non-fatal disease. For nasopharyngeal viral load with days from symptom onset (**d**) (p=0.0015), the AUC was 0.69 for fatal disease, 0.66, for severe non-fatal disease, and 0.73 for moderate, non-fatal disease. Additional statistics are shown in Table 2.

**Extended Data Fig. 3** | **Saliva viral load correlates with key cell and immunological factors in COVID-19. a-c, a**, Table shows readout for NIPALS calculation used to obtain VIP Plot in Figure 3. Factors are ordered from 1-15 showing the Root Mean prediction sum of squares (PRESS) (also depicted by graph) and the van der Voet T^2^ statistic. Percent of variation in y explained by each factor demonstrates that factor 1 explained 52.46% of y. Five factors could explain 91.47% of the variation in Y, however, these subsequent factors after factor 1 contributed increasingly less to explain Y as shown in the graph and table on the right. The table shows no statistical difference between Factor 2 which minimized the van der Voet T^2^ statistic and Factor 1 which minimized the number of VIP variables. Hence, Factor 1 was chosen as the optimal readout. **b**, heatmap depicts hierarchical clustering comparing 39 VIP immune factors between low, medium, and high viral load. Hierarchical clustering was Ward based. Measurements were normalized across saliva viral load groups. **c**, Heatmap shows comparison of Pearson correlation of lymphocytes and subsets to saliva and nasopharyngeal viral loads. Only significant correlations are shown with corresponding value inside the box.

**Extended Data Fig. 4** | **Comparison of cytokine levels amongst uninfected healthcare workers and those with low, medium and high saliva viral loads. a**, Comparison of mean cytokine levels amongst saliva viral load levels and uninfected healthcare workers. Comparisons were done using a one-way ANOVA p-values were adjusted using Tukey method for multiple comparisons.

**Extended Data Fig. 5** | **Comparison of platelets and lymphocyte subsets amongst saliva and nasopharyngeal viral load levels and amongst deceased a-c**, Individuals with saliva and nasopharyngeal viral loads were stratified into three groups based on the quantile distribution of viral load levels across the entire cohort irrespective of disease severity for all timepoints collected. For saliva these distributions were low (saliva viral load=3.212-4.4031 Log_10_ [Log](GE/ml)), medium (saliva viral load=4.4031-6.1106 Log (GE/ml)), and high (saliva viral load=6.1106-10.320 Log (GE/ml)). For nasopharyngeal viral load these measurements were low (NP viral load=3.212-4.057 Log_10_ [Log](GE/ml)), medium (NP viral load=4.057-5.76 Log (GE/ml)), and high (NP viral load=5.76-9.82 Log (GE/ml)). **a**, Comparison of mean Lymphocyte subset counts with saliva viral load levels and uninfected healthcare workers. Comparisons were done using a one-way ANOVA p-values were adjusted using Tukey method for multiple comparisons. **b**, Comparisons of nasopharyngeal viral load as described for saliva viral load in A. **c**, Comparison of lymphocyte populations as in A and B, amongst alive and deceased individuals using a 2-sided t test. Significance values were adjusted for multiple comparisons using the Holm-Sidak method.

**Extended Data Fig. 6** |**cTfh Cell kinetics amongst deceased is associated with the production of AntiRbD IgG and saliva, not nasopharyngeal, viral load. a**, Least squares means analysis comparing circulating Tfh cells in alive vs deceased individuals over days from symptom onset. Timepoints represent 10-day bins. p values are adjusted for multiple comparisons using Sidak correction. Whiskers and shading represent the 95% confidence interval of the mean. **b**, Linear Regression of Anti S1 IgG and Anti RBD IgG to saliva viral load. Lines are colored as indicated. Linear regression shows Pearson’s correlation coefficients; shading represents the 95% confidence interval for the regression line. The slopes of Anti S1 IgG was determined to be - 0.93 while the slope of Anti RBD IgG was found to be -2.0. Lines differed significantly from the average (Avg) slope which was determined to be 1.17 based on the Nelson adjusted mean (p=.0490). Dotted lines VLP=positivity threshold for viral load is 3.75 Log (Genomic Equivalents [GE]/ml)); VLD=viral load limit of detection is 3.22 Log (GE/ml). **c**, Ratio of Anti RBD IgG over saliva viral load; scatter plot showing the ratio of Anti RBD IgG over the saliva viral load of each patient timepoint as indicated. Coloring of each dot is based on saliva viral load levels as indicated on the graph. Each dot represents a patient timepoint. Lines connect repeated measurements of the same individual over time (days from symptom onset). **d**, Comparison of the mean duration of days from symptom onset in the high, medium and low nasopharyngeal viral load categories amongst spectrum of disease severity. <u>Top Panel</u> shows the Kernel projected percent distribution of time spent for individuals within severity subgroups in the high, medium, or low nasopharyngeal viral load categories. <u>Bottom panel</u> compares the mean time (y axis) of each severity subgroup within each nasopharyngeal viral load category. Points represent a patient time point. Severity is color-coded as indicated on the graph. Lines connect the means within each severity subgroup in each viral load category. Whiskers represent the standard deviation of the mean and shading represents the 95% bootstrap confidence of fit. Comparisons within each viral load category were done by one-way ANOVA and significance was adjusted for multiple comparisons across all categories using the Holm-Sidak method. The dotted line in the top and bottom panels marks day 14 from symptom onset (DfSO) which represents the rounded average of one standard deviation away from the mean for the moderate and severe groups of patients in the high saliva viral load (see Figure 4) category who survived. e, Panels are subdivided by day 14 from symptom onset as indicated in the Figure. Panels represent the frequency of individuals within each severity group that fall into the high, medium, and low nasopharyngeal viral load categories before day 14 from symptom onset (bottom left) and at or after day 14 from symptom onset (top left). f, Subpanels show the prevalence of deceased individuals within the high, medium, and low nasopharyngeal viral load categories before day 14 (bottom) and on or after day 14 (top). **g**, Panels show mean levels of Anti S1 IgG and Anti RBD IgG (top) and their ratios over nasopharyngeal viral load (bottom), amongst severity groups in the high medium and low nasopharyngeal viral load levels. X-axis denotes nasopharyngeal viral load levels as well as mean days from symptom onset (DfSO) of each severity group within each viral load level as determined in **d**. Comparisons of means amongst severity groups is made in the low nasopharyngeal viral load level via least squares mean analysis using REML to compare Anti S1 IgG and Anti RBD levels between the groups and within groups. P values are adjusted for multiple comparisons using Sidak correction.

**Extended Data Table 1: Cohort composition**

Chi-Squared Tests for significance use Poisson rates. COVID-19 Risk factors for Immunosuppression include patients with HIV, type-2 diabetes, chronic kidney disease, cirrhosis, and patients on a known immunosuppressive agent prior to admission.

**Extended Data Table 2: Logistic regression reports for saliva and nasopharyngeal viral load, and age to disease severity and mortality**

**Extended Data Table 3: GEE analysis and REML analysis of T cell counts accounting for age or with days from symptom onset**.

## References

1 Petrilli, C. M. et al.. Factors associated with hospital admission and critical illness among 5279 people with coronavirus disease 2019 in New York City: prospective cohort study. BMJ 369, m1966, doi:10.1136/bmj.m1966 (2020).

2 Zhou, F. et al.. Clinical course and risk factors for mortality of adult inpatients with COVID-19 in Wuhan, China: a retrospective cohort study. Lancet 395, 1054–1062, doi:10.1016/S0140-6736(20)30566-3 (2020).

3 Peckham, H. et al.. Male sex identified by global COVID-19 meta-analysis as a risk factor for death and ITU admission. Nat Commun 11, 6317, doi:10.1038/s41467-020-19741-6 (2020).

4 Ye, C. et al.. Impact of comorbidities on patients with COVID-19: A large retrospective study in Zhejiang, China. Journal of Medical Virology 92, 2821–2829, doi: https://doi.org/10.1002/jmv.26183 (2020).

5 Tay, M. Z., Poh, C. M., Renia, L., MacAry, P. A. & Ng, L. F. P. The trinity of COVID-19: immunity, inflammation and intervention. Nature reviews. Immunology 20, 363–374, doi:10.1038/s41577-020-0311-8 (2020).

6 Giannis, D., Ziogas, I. A. & Gianni, P. Coagulation disorders in coronavirus infected patients: COVID-19, SARS-CoV-1, MERS-CoV and lessons from the past. J Clin Virol 127, 104362, doi:10.1016/j.jcv.2020.104362 (2020).

7 Borczuk, A. C. et al.. COVID-19 pulmonary pathology: a multi-institutional autopsy cohort from Italy and New York City. Mod Pathol, doi:10.1038/s41379-020-00661-1 (2020).

8 Lucas, C. et al.. Longitudinal analyses reveal immunological misfiring in severe COVID-19. Nature 584, 463–469, doi:10.1038/s41586-020-2588-y (2020).

9 Blanco-Melo, D. et al.. Imbalanced Host Response to SARS-CoV-2 Drives Development of COVID-19. Cell 181, 1036–1045 e1039, doi:10.1016/j.cell.2020.04.026 (2020).

10 Laing, A. G. et al.. A dynamic COVID-19 immune signature includes associations with poor prognosis. Nature medicine, doi:10.1038/s41591-020-1038-6 (2020).

11 Rydyznski Moderbacher, C. et al. Antigen-Specific Adaptive Immunity to SARS-CoV-2 in Acute COVID-19 and Associations with Age and Disease Severity. Cell, doi:10.1016/j.cell.2020.09.038 (2020).

12 Garcia-Beltran, W. F. et al.. COVID-19 neutralizing antibodies predict disease severity and survival. medRxiv, 2020.2010.2015.20213512, doi:10.1101/2020.10.15.20213512 (2020).

13 Zohar, T. et al.. Compromised Humoral Functional Evolution Tracks with SARS-CoV-2 Mortality. Cell 183, 1508–1519 e1512, doi:10.1016/j.cell.2020.10.052 (2020).

14 Kaneko, N. et al.. Loss of Bcl-6-Expressing T Follicular Helper Cells and Germinal Centers in COVID-19. Cell, doi:10.1016/j.cell.2020.08.025 (2020).

15 Walsh, K. A. et al.. SARS-CoV-2 detection, viral load and infectivity over the course of an infection. J Infect 81, 357–371, doi:10.1016/j.jinf.2020.06.067 (2020).

16 Buchan, B. W. et al.. Distribution of SARS-CoV-2 PCR Cycle Threshold Values Provide Practical Insight Into Overall and Target-Specific Sensitivity Among Symptomatic Patients. Am J Clin Pathol 154, 479–485, doi:10.1093/ajcp/aqaa133 (2020).

17 Chen, C. et al.. SARS-CoV-2-Positive Sputum and Feces After Conversion of Pharyngeal Samples in Patients With COVID-19. Ann Intern Med 172, 832–834, doi:10.7326/M20-0991 (2020).

18 Barth, R. E. & De Regt, M. J. A. Persistence of viral RNA in stool samples from patients recovering from covid-19. BMJ 369, m1724, doi:10.1136/bmj.m1724 (2020).

19 Zheng, S. et al.. Viral load dynamics and disease severity in patients infected with SARS-CoV-2 in Zhejiang province, China, January-March 2020: retrospective cohort study. BMJ 369, m1443, doi:10.1136/bmj.m1443 (2020).

20 Shi, F. et al.. Association of viral load with serum biomakers among COVID-19 cases. Virology 546, 122–126, doi:10.1016/j.virol.2020.04.011 (2020).

21 Wolfel, R. et al.. Virological assessment of hospitalized patients with COVID-2019. Nature 581, 465–469, doi:10.1038/s41586-020-2196-x (2020).

22 Byrne, R. L. et al.. Saliva Alternative to Upper Respiratory Swabs for SARS-CoV-2 Diagnosis. Emerg Infect Dis 26, doi:10.3201/eid2611.203283 (2020).

23 Kim, S. E. et al.. Viral Load Kinetics of SARS-CoV-2 Infection in Saliva in Korean Patients: a Prospective Multi-center Comparative Study. J Korean Med Sci 35, e287, doi:10.3346/jkms.2020.35.e287 (2020).

24 Zhu, J., Guo, J., Xu, Y. & Chen, X. Viral dynamics of SARS-CoV-2 in saliva from infected patients. J Infect 81, e48–e50, doi:10.1016/j.jinf.2020.06.059 (2020).

25 Azzi, L. et al.. Saliva is a reliable tool to detect SARS-CoV-2. J Infect 81, e45–e50, doi:10.1016/j.jinf.2020.04.005 (2020).

26 Goel, R., Arora, R., Chhabra, M. & Kumar, S. Viral shedding in tears of COVID-19 cases presenting as conjunctivitis. Indian J Ophthalmol 68, 2308, doi:10.4103/ijo.IJO_2567_20 (2020).

27 Westblade, L. F. et al.. SARS-CoV-2 Viral Load Predicts Mortality in Patients with and without Cancer Who Are Hospitalized with COVID-19. Cancer Cell, doi:10.1016/j.ccell.2020.09.007 (2020).

28 Pujadas, E. et al.. SARS-CoV-2 viral load predicts COVID-19 mortality. Lancet Respir Med 8, e70, doi:10.1016/S2213-2600(20)30354-4 (2020).

29 Fakheran, O., Dehghannejad, M. & Khademi, A. Saliva as a diagnostic specimen for detection of SARS-CoV-2 in suspected patients: a scoping review. Infect Dis Poverty 9, 100, doi:10.1186/s40249-020-00728-w (2020).

30 Kam, K. Q. et al.. A Well Infant With Coronavirus Disease 2019 With High Viral Load. Clin Infect Dis 71, 847–849, doi:10.1093/cid/ciaa201 (2020).

31 Argyropoulos, K. V. et al.. Association of Initial Viral Load in Severe Acute Respiratory Syndrome Coronavirus 2 (SARS-CoV-2) Patients with Outcome and Symptoms. Am J Pathol 190, 1881–1887, doi:10.1016/j.ajpath.2020.07.001 (2020).

32 Jacot, D., Greub, G., Jaton, K. & Opota, O. Viral load of SARS-CoV-2 across patients and compared to other respiratory viruses. Microbes and infection / Institut Pasteur 22, 617–621, doi:10.1016/j.micinf.2020.08.004 (2020).

33 Fajnzylber, J. et al.. SARS-CoV-2 viral load is associated with increased disease severity and mortality. Nat Commun 11, 5493, doi:10.1038/s41467-020-19057-5 (2020).

34 Wyllie, A. L. et al.. Saliva or Nasopharyngeal Swab Specimens for Detection of SARS-CoV- 2. The New England journal of medicine 383, 1283–1286, doi:10.1056/NEJMc2016359 (2020).

35 To, K. K. et al.. Temporal profiles of viral load in posterior oropharyngeal saliva samples and serum antibody responses during infection by SARS-CoV-2: an observational cohort study. Lancet Infect Dis 20, 565–574, doi:10.1016/S1473-3099(20)30196-1 (2020).

36 Hamming, I. et al.. Tissue distribution of ACE2 protein, the functional receptor for SARS coronavirus. A first step in understanding SARS pathogenesis. J Pathol 203, 631–637, doi:10.1002/path.1570 (2004).

37 Xu, H. et al.. High expression of ACE2 receptor of 2019-nCoV on the epithelial cells of oral mucosa. Int J Oral Sci 12, 8, doi:10.1038/s41368-020-0074-x (2020).

38 Liu, L. et al.. Epithelial cells lining salivary gland ducts are early target cells of severe acute respiratory syndrome coronavirus infection in the upper respiratory tracts of rhesus macaques. Journal of virology 85, 4025–4030, doi:10.1128/JVI.02292-10 (2011).

39 Soares, C. D., Mosqueda-Taylor, A., Hernandez-Guerrero, J. C., de Carvalho, M. G. F. & de Almeida, O. P. Immunohistochemical expression of angiotensin-converting enzyme 2 (ACE2) in minor salivary glands during SARS-CoV-2 infection. J Med Virol, doi:10.1002/jmv.26723 (2020).

40 Dickey, B. F. What it takes for a cough to expel mucus from the airway. Proceedings of the National Academy of Sciences 115, 12340–12342, doi:10.1073/pnas.1817484115 (2018).

41 Takahashi, T. et al.. Sex differences in immune responses that underlie COVID-19 disease outcomes. Nature, doi:10.1038/s41586-020-2700-3 (2020).

42 Zhang, X. et al.. Viral and host factors related to the clinical outcome of COVID-19. Nature 583, 437–440, doi:10.1038/s41586-020-2355-0 (2020).

43 Long, Q. X. et al.. Antibody responses to SARS-CoV-2 in patients with COVID-19. Nature medicine 26, 845–848, doi:10.1038/s41591-020-0897-1 (2020).

44 Shaw, A. C., Goldstein, D. R. & Montgomery, R. R. Age-dependent dysregulation of innate immunity. Nature reviews. Immunology 13, 875–887, doi:10.1038/nri3547 (2013).

45 Smith, M., Honce, R. & Schultz-Cherry, S. Metabolic Syndrome and Viral Pathogenesis: Lessons from Influenza and Coronaviruses. Journal of virology 94, doi:10.1128/JVI.00665-20 (2020).

46 Chemaly, R. F. et al.. A multicenter study of pandemic influenza A (H1N1) infection in patients with solid tumors in 3 countries: early therapy improves outcomes. Cancer 118, 4627–4633, doi:10.1002/cncr.27447 (2012).

47 Pinto, B. G. G. et al.. ACE2 Expression Is Increased in the Lungs of Patients With Comorbidities Associated With Severe COVID-19. Journal of Infectious Diseases 222, 556–563, doi:10.1093/infdis/jiaa332 (2020).

48 Wang, Y. et al.. Remdesivir in adults with severe COVID-19: a randomised, double-blind, placebo-controlled, multicentre trial. Lancet 395, 1569–1578, doi:10.1016/S0140-6736(20)31022-9 (2020).

49 Lyngbakken, M. N. et al.. A pragmatic randomized controlled trial reports lack of efficacy of hydroxychloroquine on coronavirus disease 2019 viral kinetics. Nat Commun 11, 5284, doi:10.1038/s41467-020-19056-6 (2020).

50 Company, E. L. a. Lilly Statement Regarding NIH’s ACTIV-3 Clinical Trial, https://www.lilly.com/news/stories/statement-activ3-clinical-trial-nih-covid19 (2020).

51 Regeneron. REGN-COV2 INDEPENDENT DATA MONITORING COMMITTEE RECOMMENDS HOLDING ENROLLMENT IN HOSPITALIZED PATIENTS WITH HIGH OXYGEN REQUIREMENTS AND CONTINUING ENROLLMENT IN PATIENTS WITH LOW OR NO OXYGEN REQUIREMENTS, https://newsroom.regeneron.com/news-releases/news-release-details/regn-cov2-independent-data-monitoring-committee-recommends (2020).

52 Vogels, C. B. F. et al.. Analytical sensitivity and efficiency comparisons of SARS-CoV-2 RT- qPCR primer-probe sets. Nat Microbiol 5, 1299–1305, doi:10.1038/s41564-020-0761-6 (2020).

53 Amanat, F. et al.. A serological assay to detect SARS-CoV-2 seroconversion in humans. Nature medicine 26, 1033–1036, doi:10.1038/s41591-020-0913-5 (2020).

