## Extended Data Figures 1-6 for "Saliva viral load is a dynamic unifying correlate of COVID-19 severity and mortality"

### Extended Figure 1

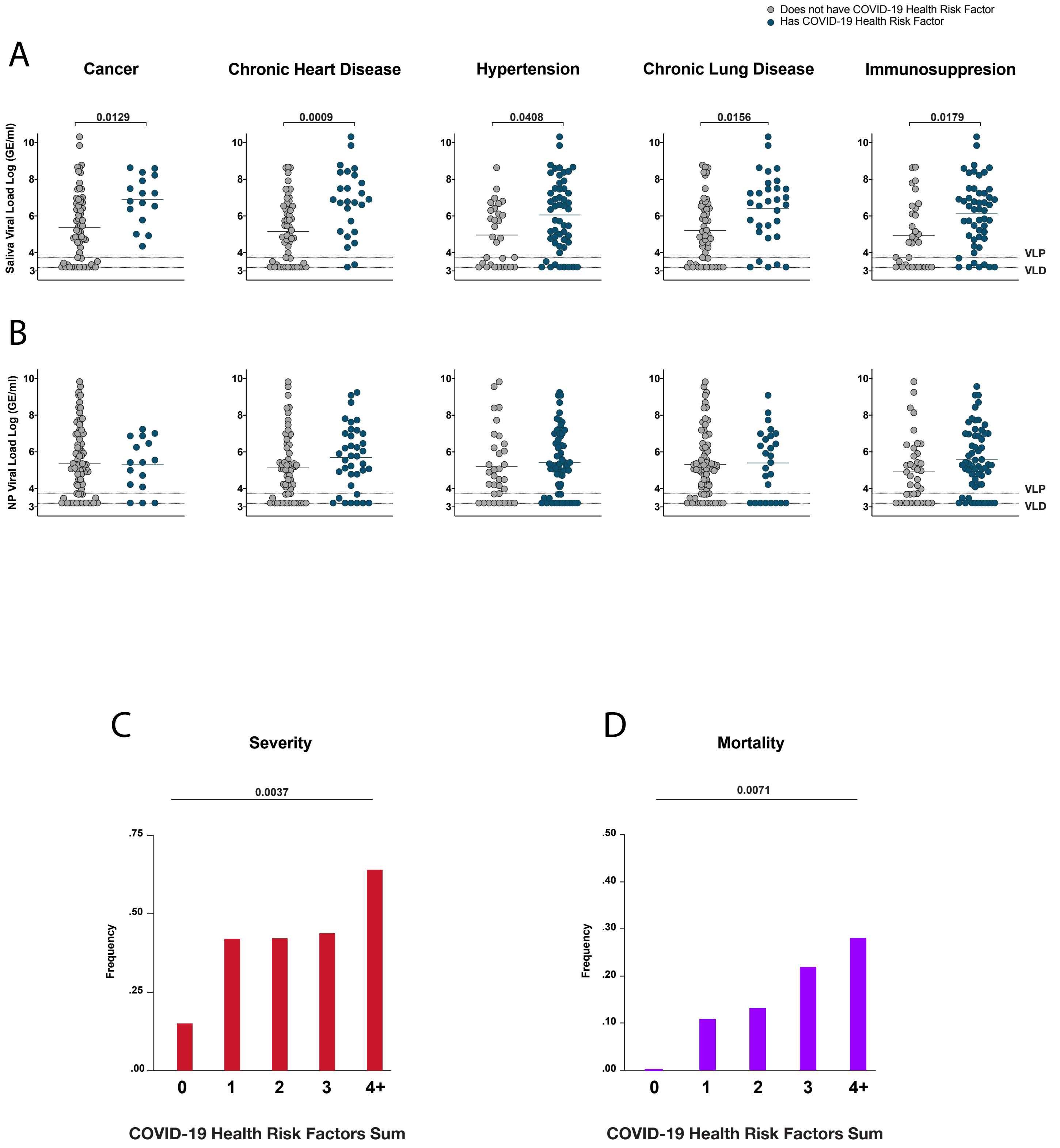

### Extended Figure 2:

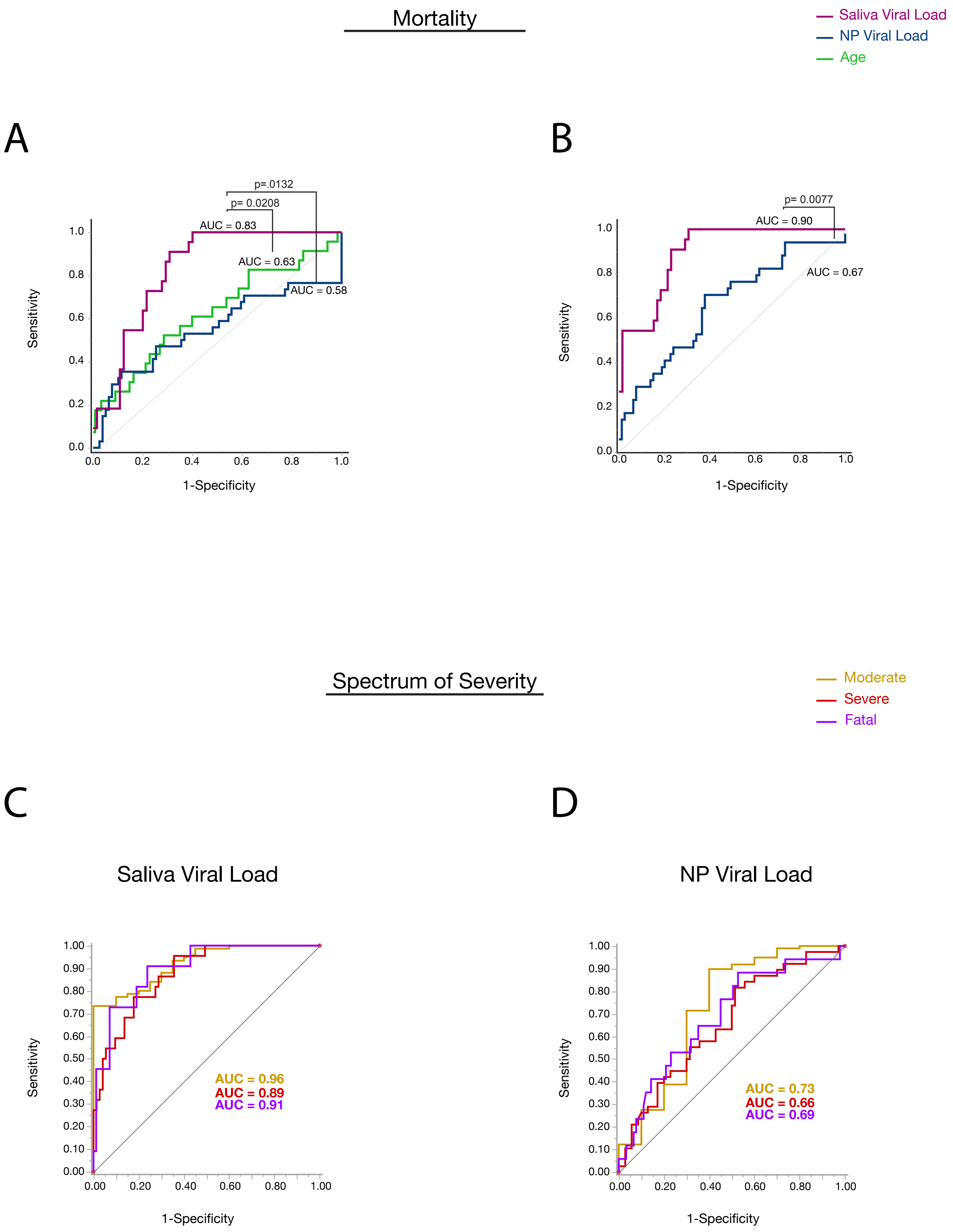

### Extended Figure 3

A

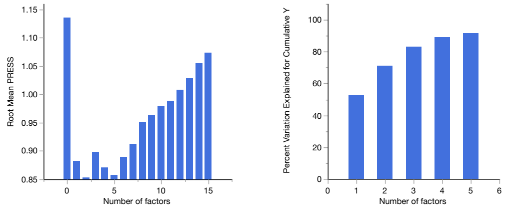


| Number of factors | Root Mean PRESS | van der Voet $T^2$ | Prob > van der Voet $T^2$ | $Q^2$ | Cumulative $Q^2$ | $R^2X$ | Cumulative $R^2X$ | $R^2Y$ | Cumulative $R^2Y$ | Method | Number of rows | Percent Variation Explained for Cumulative X | Percent Variation Explained for Cumulative Y | Number of VIP > 0.8 |
|---|---|---|---|---|---|---|---|---|---|---|---|---|---|---|
| 0 | 1.135 | 7.688 | 0.0060* | -0.120 | -0.120 | 0 | 0 | 0 | 0 | | | | | |
| **1** | **0.882** | **0.241** | **0.645** | **0.317** | **0.317** | **0.169** | **0.169** | **0.546** | **0.546** | **NIPALS** | **59** | **16.59** | **52.46** | **39** |
| 2 | 0.853 | 0.000 | 1 | 0.359 | 0.562 | 0.091 | 0.260 | 0.202 | 0.748 | NIPALS | 59 | 25.64 | 71.08 | 50 |
| 3 | 0.898 | 1.676 | 0.206 | 0.292 | 0.690 | 0.076 | 0.336 | 0.111 | 0.859 | NIPALS | 59 | 32.94 | 82.86 | 50 |
| 4 | 0.871 | 0.131 | 0.734 | 0.335 | 0.794 | 0.049 | 0.385 | 0.055 | 0.915 | NIPALS | 59 | 37.48 | 89.09 | 50 |
| 5 | 0.858 | 0.004 | 0.94 | 0.353 | 0.866 | 0.044 | 0.430 | 0.025 | 0.940 | NIPALS | 59 | 41.62 | 91.47 | 50 |
| 6 | 0.890 | 0.308 | 0.578 | 0.304 | 0.907 | 0.040 | 0.470 | 0.017 | 0.957 | | | | | |
| 7 | 0.913 | 0.715 | 0.429 | 0.268 | 0.932 | 0.037 | 0.508 | 0.011 | 0.968 | | | | | |
| 8 | 0.951 | 1.761 | 0.173 | 0.204 | 0.946 | 0.042 | 0.550 | 0.007 | 0.975 | | | | | |
| 9 | 0.963 | 1.884 | 0.173 | 0.183 | 0.956 | 0.034 | 0.584 | 0.006 | 0.981 | | | | | |
| 10 | 0.980 | 2.256 | 0.13 | 0.154 | 0.963 | 0.026 | 0.610 | 0.005 | 0.986 | | | | | |
| 11 | 0.989 | 2.327 | 0.138 | 0.138 | 0.968 | 0.024 | 0.634 | 0.004 | 0.990 | | | | | |
| 12 | 1.008 | 2.742 | 0.101 | 0.105 | 0.971 | 0.031 | 0.665 | 0.002 | 0.992 | | | | | |
| 13 | 1.029 | 3.324 | 0.07 | 0.067 | 0.973 | 0.023 | 0.688 | 0.002 | 0.994 | | | | | |
| 14 | 1.055 | 4.051 | 0.0350* | 0.018 | 0.974 | 0.021 | 0.708 | 0.001 | 0.996 | | | | | |
| 15 | 1.074 | 4.605 | 0.0200* | -0.017 | 0.973 | 0.018 | 0.726 | 0.001 | 0.997 | | | | | |

**KFold Cross Validation with K=5**

B

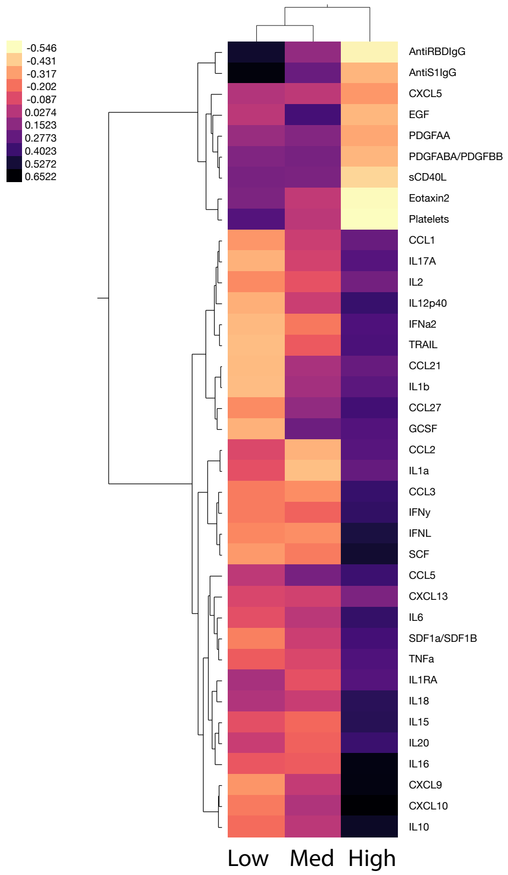


C

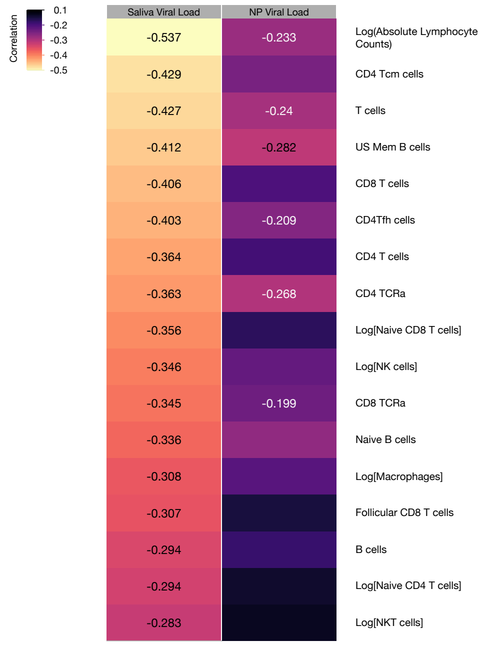

### Extended Figure 4

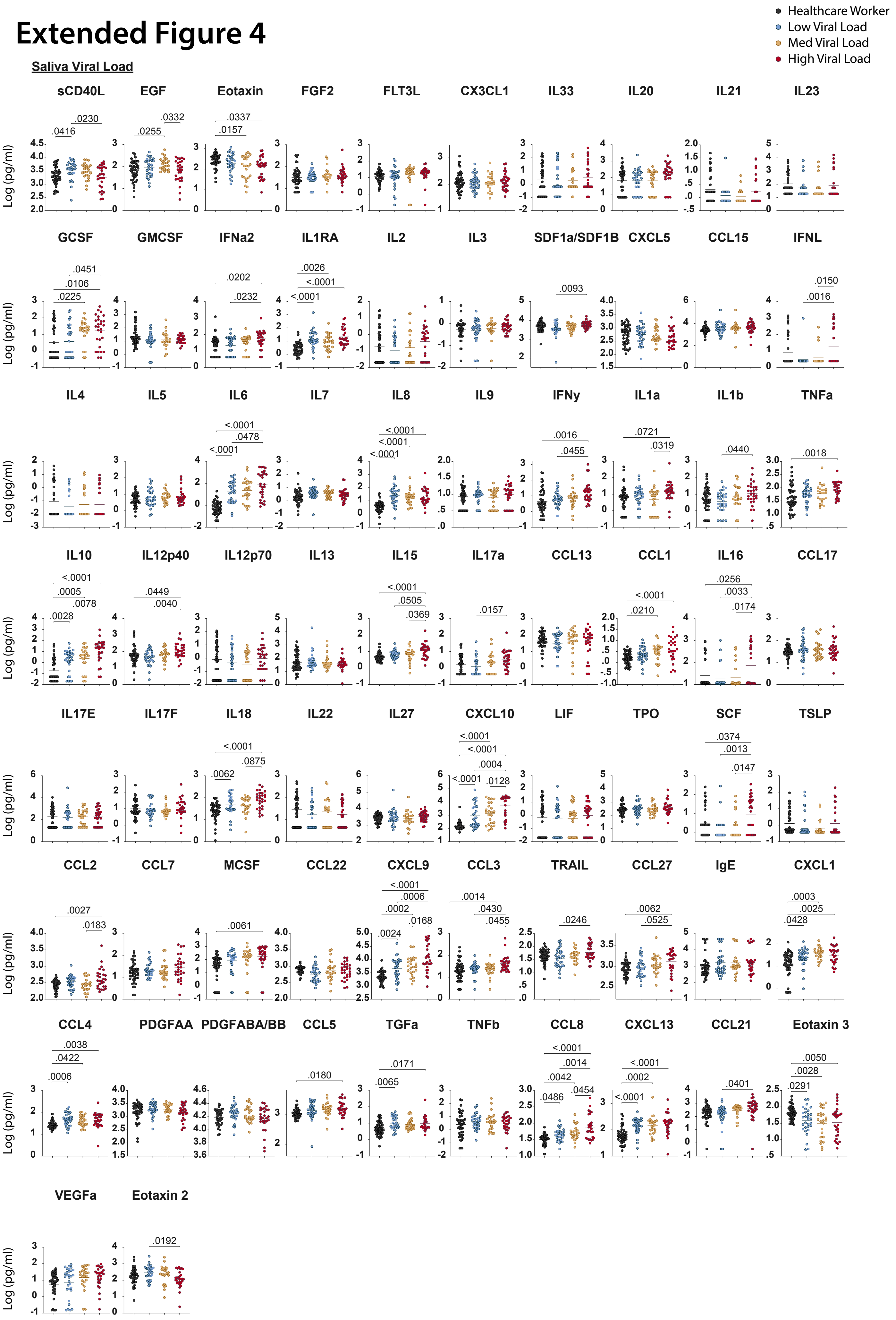

### Extended Figure 5

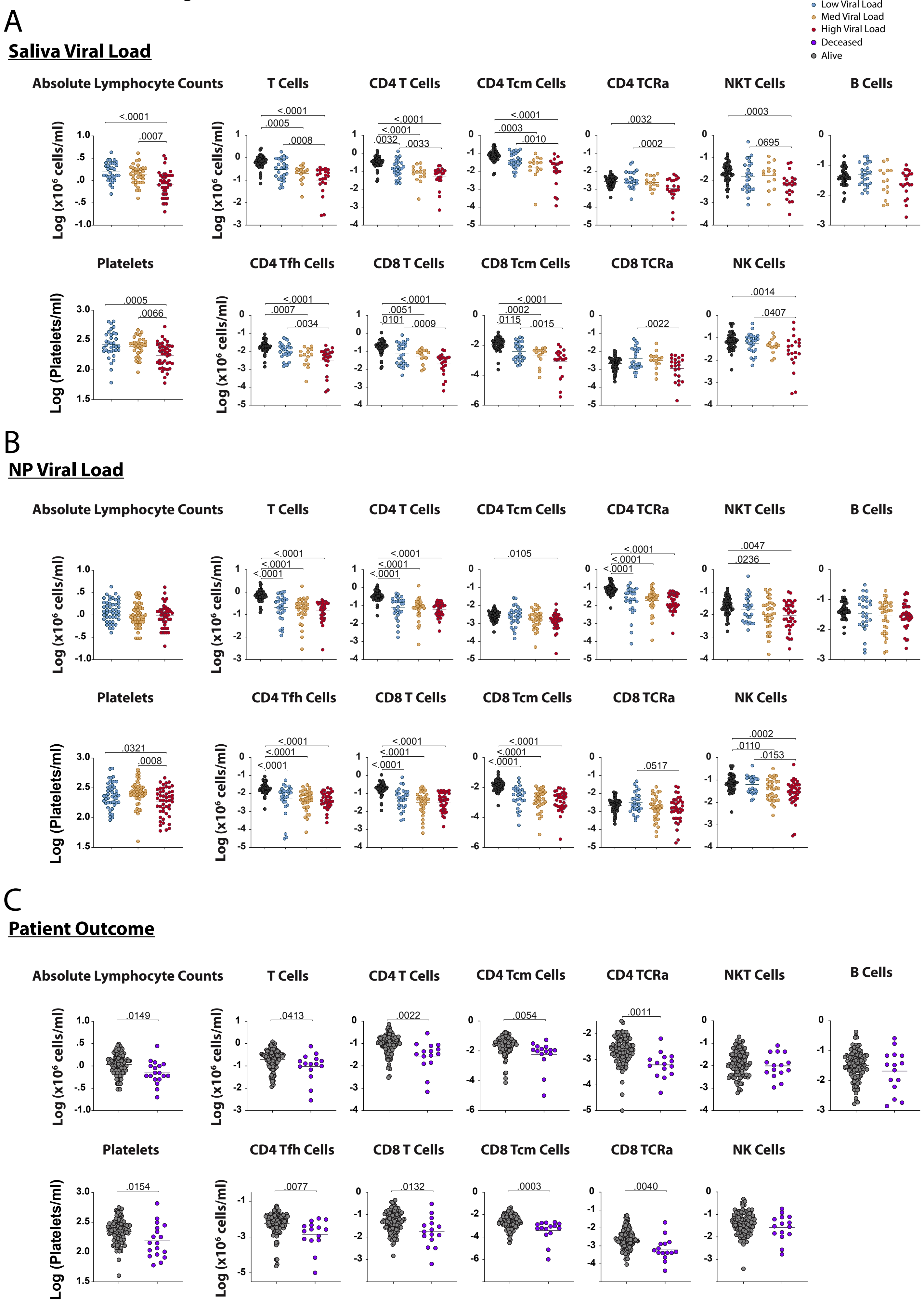

### Extended Figure 6:

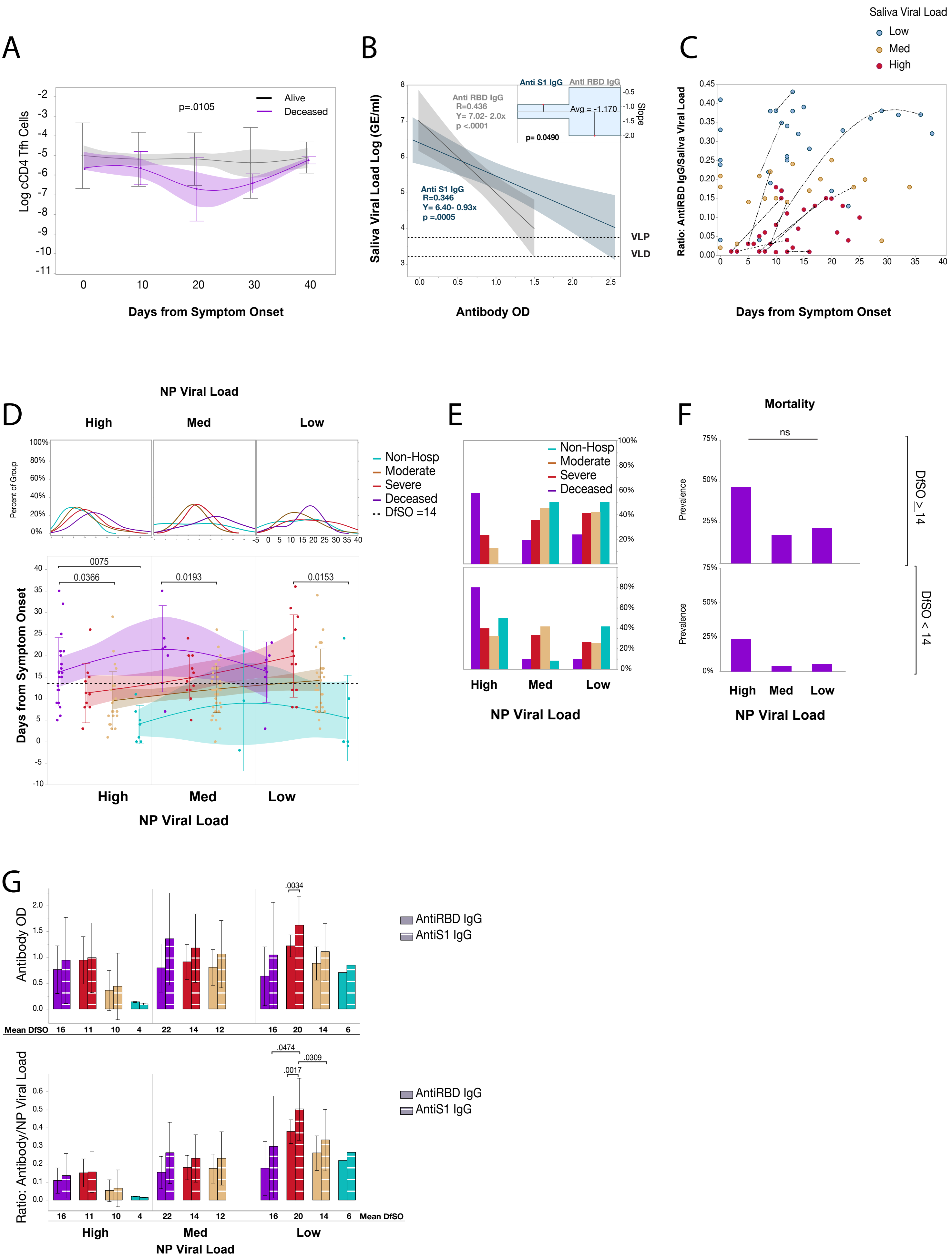

### Extended Figure 7: Gating Strategies for cell populations

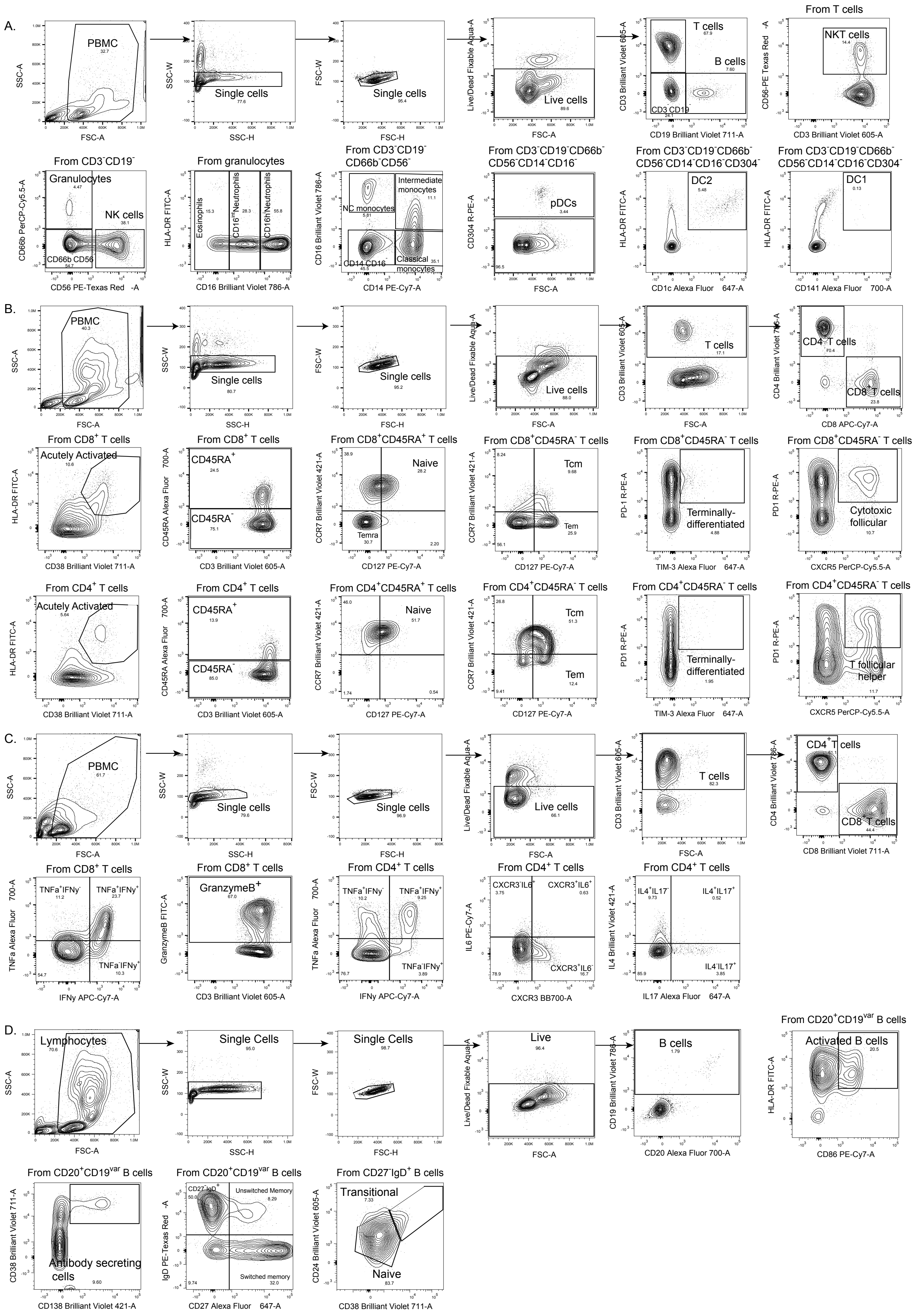
