## Supplemental Tables for "Saliva viral load is a dynamic unifying correlate of COVID-19 severity and mortality"

### Table 1: Cohort Demographics

| | Complete Cohort Demographics | | | | | | Disease Severity Amongst Patients Hospitalized for COVID-19 | | | | Patient Outcome | | | |
|---|---|---|---|---|---|---|---|---|---|---|---|---|---|---|
| | Uninfected | Infected | | | | | | | | | | | | |
| | Healthcare Workers | Not Hospitalized for COVID-19 | Moderate Disease | Severe Disease | Fatal COVID-19 | Total Cohort | Moderate | Severe | | All | Alive | Deceased | | All |
| | N (% Total) | N (% Total) | N (% Total) | N (% Total) | N (% Total) | N (% Total) | N (% Total) | N (% Total) | | N (% Total) | N (% Total) | N (% Total) | | N (% Total) |
| Summary | 109 (38.79%) | 26 (9.25%) | 81 (28.83%) | 42 (14.95%) | 23 (8.19%) | 281 (100%) | 84 (57.53%) | 62 (42.47%) | | 146 (100%) | 131 (85.06%) | 23 (14.94%) | | 154 (100%) |
| | Mean, Std Dev | Mean, Std Dev | Mean, Std Dev | Mean, Std Dev | Mean, Std Dev | Mean, Std Dev | Mean, Std Dev | Mean, Std Dev | | Mean, Std Dev | Mean, Std Dev | Mean, Std Dev | | Mean, Std Dev |
| Age | 37.39, ±11.7 | 37.31, ±14.24 | 63.32, ±15.99 | 64.57, ±14.47 | 71.09, ±18.34 | 51.68, ±19.87 | 64.12, ±16.40 | 65.89, ±15.7 | | 64.9, ±16.09 | 62, ±16.68 | 71.09, ±18.34 | | 63.36, ±17.18 |
| Age Group | N (%) | N (%) | N (%) | N (%) | N (%) | N (%) | N (%) | N (%) | Relative Risk (95% CI), **p-value*** | N (%) | N (%) | N (%) | Relative Risk (95% CI), **p-value*** | |
| <59 | 105 (96.33%) | 22 (84.62%) | 30 (37.04%) | 12 (28.57%) | 4 (17.39%) | 173 (61.57%) | 30 (35.71%) | 16 (25.81%) | 0.67 (0.39-1.15) | 46 (31.51%) | 50 (38.17%) | 4 (17.39%) | 0.48 (0.19-1.19) | 54 (35.06%) |
| 59-74 | 4 (3.67%) | 4 (15.38%) | 28 (34.57%) | 17 (40.48%) | 7 (30.43%) | 60 (21.35%) | 28 (33.33%) | 24 (38.71%) | 1.27 (0.81-1.99) | 52 (35.62%) | 45 (34.35%) | 7 (30.43%) | 0.8 (0.39-1.65) | 52 (33.77%) |
| >74 | 0 (0%) | 0 (0%) | 23 (28.4%) | 13 (30.95%) | 12 (52.17%) | 48 (17.08%) | 26 (30.95%) | 22 (35.48%) | 1.11 (0.69-1.78) | 48 (32.88%) | 36 (27.48%) | 12 (52.17%) | 1.96 (1.22-3.15) | 48 (31.17%) |
| Sex | | | | | | | | | | | | | | |
| F | 83 (76.15%) | 22 (84.62%) | 39 (48.15%) | 21 (50%) | 8 (34.78%) | 173 (61.57%) | 41 (48.81%) | 27 (43.55%) | 0.80 (0.56-1.16) | 68 (46.58%) | 66 (50.38%) | 8 (34.78%) | 0.73 (0.41-1.3) | 74 (48.05%) |
| M | 26 (23.85%) | 4 (15.38%) | 42 (51.85%) | 21 (50%) | 15 (65.22%) | 108 (38.43%) | 43 (51.19%) | 35 (56.45%) | .86 (0.83-1.53) | 78 (53.42%) | 65 (49.62%) | 15 (65.22%) | 1.27 (0.89-1.83) | 80 (51.95%) |
| Race or Ethnicity | | | | | | | | | | | | | | |
| American Indian/ Alaskan native | 0 (0%) | | | | | | | | – | | | | – | |
| Asian | 7 (6.42%) | 1 (3.85%) | 1 (1.08%) | 0 (0%) | 1 (4.35%) | 10 (3.56%) | 0 (0%) | 3 (4.84%) | – | 2 (1.37%) | 1 (0.76%) | 1 (4.35%) | – | 2 (1.3%) |
| Black/African American [Not Hispanic/Latin(a/o)] | 6 (5.5%) | 3 (11.54%) | 24 (29.63%) | 15 (50%) | 7 (30.43%) | 60 (21.35%) | 26 (30.95%) | 25 (40.32%) | 1.33 (0.86-2.06) | 51 (34.93%) | 46 (35.11%) | 7 (30.43%) | 0.87 (0.45-1.68) | 53 (34.42%) |
| Native Hawaiian/ Pacific Islander | 0 (0%) | 0 (0%) | 0 (0%) | 0 (0%) | 0 (0%) | 0 (0%) | 0 (0%) | 0 (0%) | – | 0 (0%) | 0 (0%) | 0 (0%) | – | 0 (0%) |
| White/Caucasian [Not Hispanic/Latin(a/o)] | 82 (75.23%) | 18 (69.23%) | 42 (51.85%) | 11 (36.67%) | 11 (47.83%) | 168 (59.79%) | 43 (51.19%) | 24 (38.71%) | 0.80 (0.56-1.16) | 68 (46.58%) | 59 (45.04%) | 11 (47.83%) | 1.09 (0.69-1.72) | 70 (45.45%) |
| Hispanic/Latin(a/o) of any race | 8 (7.34%) | 4 (15.38%) | 13 (16.05%) | 4 (13.33%) | 4 (17.39%) | 34 (12.10%) | 13 (15.48%) | 9 (14.52%) | 1.00 (0.46-2.17) | 22 (15.07%) | 22 (16.79%) | 4 (17.39%) | 1.04 (0.4-2.73) | 26 (16.88%) |
| Multiple | 0 (0%) | 0 (0%) | 0 (0%) | 0 (0%) | 0 (0%) | 0 (0%) | 0 (0%) | 0 (0%) | – | 0 (0%) | 0 (0%) | 0 (0%) | – | 0 (0%) |
| Unknown | 6 (5.5%) | 0 (0%) | 2 (2.47%) | 0 (0%) | 0 (0%) | 9 (3.20%) | 2 (2.38%) | 1 (1.61%) | – | 3 (2.05%) | 3 (2.29%) | 0 (0%) | – | 3 (1.95%) |
| HCW/Patient Breakdown | | | | | | | | | | | | | | |
| HCW | 109 (100%) | 18 (69.23%) | 0 (0%) | 0 (0%) | 0 (0%) | 127 (45.20%) | 0 (0%) | 0 (0%) | | 0 (0%) | 0 (0%) | 0 (0%) | | 0 (0%) |
| Pateint | 0 (0%) | 8 (30.77%) | 81 (100%) | 42 (100%) | 23 (100%) | 154 (54.80%) | 84 (100%) | 62 (100%) | | 146 (100%) | 131 (100%) | 23 (100%) | | 154 (100%) |
| Mortality | | | | | | | 3 (3.57%) | 20 (32.26%) | 8.72 (2.70-28.16), **<.0001** | | | | | |
| Severity | | | | | | | | | | | | | | |
| MODERATE | | | | | | | | | | | | 5 (21.74%) | 0.29 (0.13-0.63), **.001** | |
| SEVERE | | | | | | | | | | | | 18 (78.26%) | 3.21 (2.2-4.68), **.0003** | |
| Health Risk Factors Amongst Patients | | | | | | | | | | | | | | |
| Cancer within 1 year | | 0 (0%) | 11 (13.58%) | 4 (9.52%) | 5 (21.74%) | 20 (12.99%) | 12 (14.29%) | 8 (12.9%) | 0.92 (0.40-2.11) | 20 (13.7%) | 15 (11.45%) | 5 (21.74%) | 2.05 (0.82-5.11) | 20 (12.99%) |
| Chronic Heart Disease (CHD) | | 2 (25%) | 24 (29.63%) | 18 (42.86%) | 12 (52.17%) | 56 (36.36%) | 25 (29.76%) | 29 (46.77%) | 1.54 (1.01-2.36) | 54 (36.99%) | 44 (33.59%) | 12 (52.17%) | 1.5 (0.92-2.44) | 56 (36.36%) |
| Hypertension (HTN) | | 2 (25%) | 46 (56.79%) | 31 (73.81%) | 20 (86.96%) | 99 (64.29%) | 48 (57.14%) | 49 (79.03%) | 1.38 (1.10-1.73) | 97 (66.44%) | 79 (60.31%) | 20 (86.96%) | 1.45 (1.16-1.81) | 99 (64.29%) |
| Chronic Lung Disease | | 1 (12.5%) | 25 (30.86%) | 17 (40.48%) | 9 (39.13%) | 52 (33.77%) | 25 (29.76%) | 26 (41.94%) | 1.38 (0.88-2.15) | 51 (34.93%) | 43 (32.82%) | 9 (39.13%) | 1.09 (0.6-2.0) | 52 (33.77%) |
| Immunosuppression* | | 1 (12.5%) | 43 (53.09%) | 25 (59.52%) | 17 (73.91%) | 86 (55.84%) | 45 (53.57%) | 40 (64.52%) | 1.19 (0.91-1.57) | 85 (58.22%) | 69 (52.67%) | 17 (73.91%) | 1.39 (1.02-1.88) | 86 (55.84%) |
| COVID-19 HEALTH RISK FACTORS SUM | | | | | | | | | | | | | | |
| 0 | | 2 (25%) | 17 (20.99%) | 3 (7.14%) | 0 (0%) | 22 (14.29%) | 17 (20.24%) | 3 (4.84%) | **0.24 (0.07-0.79), .0089** | 20 (13.7%) | 22 (16.79%) | 0 (0%) | 0 (0-0) | 22 (14.29%) |
| 1 | | 6 (75%) | 17 (20.99%) | 10 (23.81%) | 4 (17.39%) | 37 (24.03%) | 18 (21.43%) | 13 (20.97%) | 0.99 (0.53-1.87) | 31 (21.23%) | 33 (25.19%) | 4 (17.39%) | 0.74 (0.29-1.89) | 37 (24.03%) |
| 2 | | 0 (0%) | 21 (25.93%) | 12 (28.57%) | 5 (21.74%) | 38 (24.68%) | 22 (26.19%) | 16 (25.81%) | 1.00 (0.58-1.74) | 38 (26.03%) | 33 (25.19%) | 5 (21.74%) | 0.89 (0.39-2.05) | 38 (24.68%) |
| 3 | | 0 (0%) | 17 (20.99%) | 8 (19.05%) | 7 (30.43%) | 32 (20.78%) | 18 (21.43%) | 14 (22.58%) | 1.07 (0.58-1.98) | 32 (21.92%) | 25 (19.08%) | 7 (30.43%) | 1.67 (0.82-3.4) | 32 (20.78%) |
| 4+ | | 0 (0%) | 9 (11.11%) | 9 (21.43%) | 7 (30.43%) | 25 (16.23%) | 9 (10.71%) | 16 (25.81%) | **2.30 (1.08-4.90), .0442** | 25 (17.12%) | 18 (13.74%) | 7 (30.43%) | 2.02 (0.9-4.56) | 25 (16.23%) |

### Table 2: Logistic Regression Reports

| | Predictor | Odds Ratio (per unit) | Lower 95% | Upper 95% | p value | AUC |
|---|---|---|---|---|---|---|
| Disease Severity | | | | | | |
| | Saliva Viral Load | 1.58 | 1.17 | 2.13 | 0.0012 | 0.71 |
| | NP Viral Load | 1.01 | 0.80 | 1.28 | ns | 0.51 |
| | Age | 1.00 | 0.98 | 1.02 | ns | 0.51 |
| | Days from Symptom Onset | 1.03 | 0.98 | 1.09 | ns | 0.58 |
| With Days from Symptom Onset | | | | | | |
| | Saliva Viral Load and Days from Symptom Onset | | | | <.0001* | 0.79 |
| | Saliva Viral Load | 2.03 | 1.36 | 3.03 | <.0001* | |
| | Days from Symptom Onset | 1.11 | 1.03 | 1.20 | 0.0021* | |
| | NP Viral Load and Days from Symptom Onset | | | | 0.0144 | 0.68 |
| | NP Viral Load | 1.17 | 0.89 | 1.54 | ns | |
| | Days from Symptom Onset | 1.10 | 1.03 | 1.18 | 0.0036* | |
| Mortality | | | | | | |
| | Saliva Viral Load | 2.21 | 1.35 | 3.60 | 0.0001* | 0.83 |
| | NP Viral Load | 1.23 | 0.92 | 1.64 | ns | 0.58 |
| | Age | 1.03 | 1.00 | 1.07 | 0.0376* | 0.63 |
| | Days from Symptom Onset | 1.02 | 0.96 | 1.09 | ns | 0.55 |
| With Days from Symptom Onset | | | | | | |
| | Saliva Viral Load and Days from Symptom Onset | | | | <.0001* | 0.90 |
| | Saliva Viral Load | 3.69 | 1.69 | 8.04 | <.0001* | |
| | Days from Symptom Onset | 1.18 | 1.05 | 1.33 | 0.0011* | |
| | NP Viral Load and Days from Symptom Onset | | | | 0.0619 | 0.67 |
| | NP Viral Load | 1.40 | 1.00 | 1.95 | 0.0452* | |
| | Days from Symptom Onset | 1.08 | 1.00 | 1.16 | 0.0560 | |
| Full Spectrum of Disease | | Likelihood Ratio | | | | |
| | Saliva Viral Load and Days from Symptom Onset | 40.12 | | | <.0001* | |
| | Saliva Viral Load | 39.04 | | | <.0001* | |
| | Days from Symptom Onset | 67.78 | | | <.0001* | |
| | Saliva Viral Load * Days from Symptom Onset | 9.96 | | | 0.0016 | |
| | Non-Hospitalized | | | | | — |
| | Moderate | | | | | 0.96 |
| | Severe | | | | | 0.89 |
| | Fatal | | | | | 0.91 |
| | NP Viral Load and Days from Symptom Onset | 7.69 | | | 0.0015 | |
| | NP Viral Load | 4.65 | | | 0.0311 | |
| | Days from Symptom Onset | 14.90 | | | 0.0001 | |
| | NP Viral Load * Days from Symptom Onset | 1.12 | | | ns | |
| | Non-Hospitalized | | | | | — |
| | Moderate | | | | | 0.75 |
| | Severe | | | | | 0.66 |
| | Fatal | | | | | 0.67 |

### Extended Table 3

| Term | | Least Squares Mean Difference, (95% CI) | p value | Least Squares Mean Difference, (95% CI) | p value | RESTRICTED MAXIMUM LIKLIHOOD | Estimate | R adjusted | RMSE | p value | RESTRICTED MAXIMUM LIKLIHOOD | Estimate | R adjusted | RMSE | p value |
|---|---|---|---|---|---|---|---|---|---|---|---|---|---|---|---|
| T cells | ** | Saliva Viral Load | | NP Viral Load | | * Saliva Viral Load | | 0.957 | 0.162 | 0.0024 | NP Viral Load | | 0.96 | 0.121 | 0.0361 |
| | Age | | 0.0076 | | 0.0047 | Saliva Viral Load | 0.0076, (-0.0047) | | | 0.0021 | NP Viral Load | -0.035, (-0.061--0.008) | | | 0.011 |
| | Viral Load Levels | | 0.0007 | | 0.0012 | Days from Symptom Onset | 0.0007, (-0.0012) | | | 0.0184 | Days from Symptom Onset | -0.002, (-0.009-0.005) | | | 0.512 |
| | HCW vs High | 1.3 (0.7-1.91) | <.0001 | 0.97, (0.44-1.51) | 0.0004 | | | | | | | | | | |
| | Low vs High | 0.89 (0.34-1.43) | 0.0015 | 0.2, (-0.3-0.7) | 0.440 | | | | | | | | | | |
| | Med vs High | 0.61 (-0.1-1.23) | 0.052 | 0.02, (-0.45-0.49) | 0.943 | | | | | | | | | | |
| CD8 T cells | | | | | | | | 0.964 | 0.061 | 0.0067 | | | 0.969 | 0.042 | 0.1472 |
| | Age | | 0.0011 | | 0.0036 | Saliva Viral Load | 0.0011, (-0.0036) | | | 0.0027 | NP Viral Load | -0.009, (-0.019-0) | | | 0.055 |
| | Viral Load Levels | | 0.007 | | 0.0034 | Days from Symptom Onset | 0.007, (-0.0034) | | | 0.0471 | Days from Symptom Onset | 0, (-0.003-0.002) | | | 0.791 |
| | HCW vs High | 1.27 (0.54-2.1) | 0.0006 | 0.97, (0.33-1.6) | 0.003 | | | | | | | | | | |
| | Low vs High | 0.97 (0.3-1.64) | 0.0046 | 0.24, (-0.36-0.84) | 0.430 | | | | | | | | | | |
| | Med vs High | 0.89 (0.13-1.64) | 0.0212 | -0.15, (-0.72-0.41) | 0.589 | | | | | | | | | | |
| CD8 Tcm cells | | | | | | | | 0.979 | 0.003 | 0.0001 | | | 0.959 | 0.003 | 0.0558 |
| | Age | | 0.0735 | | 0.0488 | Saliva Viral Load | 0.0735, (-0.0488) | | | 0.0002 | NP Viral Load | -0.001, (-0.001-0) | | | 0.017 |
| | Viral Load Levels | | 0.0004 | | <.0001 | Days from Symptom Onset | 0.0004, (-<.0001) | | | 0.0002 | Days from Symptom Onset | 0, (0-0) | | | 0.486 |
| | HCW vs High | 2.1 (1.16-3.5) | <.0001 | 1.73, (0.95-2.51) | <.0001 | | | | | | | | | | |
| | Low vs High | 1.39 (0.51-2.26) | 0.0019 | 0.46, (-0.28-1.2) | 0.223 | | | | | | | | | | |
| | Med vs High | 0.81 (-0.17-1.78) | 0.1050 | 0.03, (-0.66-0.72) | 0.935 | | | | | | | | | | |
| CD4 T cells | | | | | | | | 0.989 | 0.062 | 0.0271 | | | 0.984 | 0.048 | 0.0956 |
| | Age | | 0.0159 | | 0.0077 | Saliva Viral Load | 0.0159, (-0.0077) | | | 0.0133 | NP Viral Load | -0.012, (-0.024--0.001) | | | 0.037 |
| | Viral Load Levels | | 0.0027 | | 0.0004 | Days from Symptom Onset | 0.0027, (-0.0004) | | | 0.022 | Days from Symptom Onset | 0, (-0.003-0.003) | | | 0.893 |
| | HCW vs High | 1.28 (0.61-1.95) | 0.0002 | 1.07, (0.48-1.66) | 0.0004 | | | | | | | | | | |
| | Low vs High | 0.83 (0.21-1.44) | 0.0087 | 0.05, (-0.5-0.61) | 0.8469 | | | | | | | | | | |
| | Med vs High | 0.33 (-0.37-1.3) | 0.3525 | -0.03, (-0.55-0.49) | 0.9076 | | | | | | | | | | |
| CD4 Tcm cells | | | | | | | | 0.947 | 0.02 | p<.0001 | | | 0.96 | 0.015 | 0.0091 |
| | Age | | 0.206 | | 0.0769 | Saliva Viral Load | 0.206, (-0.0769) | | | 0.0003 | NP Viral Load | -0.005, (-0.009--0.002) | | | 0.002 |
| | Viral Load Levels | | 0.0009 | | 0.0046 | Days from Symptom Onset | 0.0009, (-0.0046) | | | 0.0003 | Days from Symptom Onset | 0, (-0.001-0) | | | 0.347 |
| | HCW vs High | 1.6 (0.79-2.4) | <.0001 | 1.11, (0.41-1.81) | 0.0019 | | | | | | | | | | |
| | Low vs High | 1.25 (0.5-1.99) | 0.0010 | 0.03, (-0.63-0.7) | 0.9199 | | | | | | | | | | |
| | Med vs High | 0.47 (-0.36-1.3) | 0.2645 | 0.12, (-0.5-0.73) | 0.7142 | | | | | | | | | | |
| CD4 Tfh cells | | | | | | | | 0.969 | 0.005 | 0.0004 | | | 0.957 | 0.004 | 0.0174 |
| | Age | | 0.0558 | | 0.0101 | Saliva Viral Load | 0.0558, (-0.0101) | | | 0.0009 | NP Viral Load | -0.001, (-0.002-0) | | | 0.005 |
| | Viral Load Levels | | 0.0094 | | 0.0155 | Days from Symptom Onset | 0.0094, (-0.0155) | | | 0.0009 | Days from Symptom Onset | 0, (0-0) | | | 0.423 |
| | HCW vs High | 1.18 (0.45-1.92) | 0.0016 | 0.88, (0.21-1.55) | 0.0099 | | | | | | | | | | |
| | Low vs High | 0.96 (0.28-1.63) | 0.0052 | 0.03, (-0.6-0.66) | 0.9283 | | | | | | | | | | |
| | Med vs High | 0.33 (-0.42-1.9) | 0.3864 | -0.08, (-0.67-0.51) | 0.7801 | | | | | | | | | | |
| CD4 TCRA cells | | | | | | | | 0.94 | 0.003 | 0.0219 | | | 0.981 | 0.001 | 0.0026 |
| | Age | | 0.1573 | | 0.1958 | Saliva Viral Load | 0.1573, (-0.1958) | | | 0.0077 | NP Viral Load | -0.001, (-0.001-0) | | | 0.001 |
| | Viral Load Levels | | 0.0042 | | 0.2048 | Days from Symptom Onset | 0.0042, (-0.2048) | | | 0.1563 | Days from Symptom Onset | 0, (0-0) | | | 0.876 |
| | HCW vs High | 0.49 (-0.16-1.14) | 0.1408 | 0.35, (-0.21-0.91) | 0.2249 | | | | | | | | | | |
| | Low vs High | 1.7 (0.48-1.66) | 0.0004 | 0.58, (0.05-1.11) | 0.0315 | | | | | | | | | | |
| | Med vs High | 0.72 (0.4-1.4) | 0.0391 | 0.26, (-0.24-0.76) | 0.3089 | | | | | | | | | | |
| CD8 TCRA cells | | | | | | | | 0.923 | 0.005 | 0.0668 | | | 0.949 | 0.004 | 0.0672 |
| | Age | | 0.0178 | | 0.0105 | Saliva Viral Load | 0.0178, (-0.0105) | | | 0.0206 | NP Viral Load | -0.001, (-0.002-0) | | | 0.043 |
| | Viral Load Levels | | 0.0011 | | 0.0476 | Days from Symptom Onset | 0.0011, (-0.0476) | | | 0.806 | Days from Symptom Onset | 0, (0-0) | | | 0.632 |
| | HCW vs High | -0.6 (-0.84-0.71) | 0.8770 | -0.22, (-0.94-0.5) | 0.5534 | | | | | | | | | | |
| | Low vs High | 1.2 (0.31-1.73) | 0.0047 | 0.72, (0.04-1.4) | 0.0386 | | | | | | | | | | |
| | Med vs High | 0.92 (0.12-1.72) | 0.0235 | 0.03, (-0.61-0.67) | 0.9229 | | | | | | | | | | |
| Absolute Lymphocyte Counts | | | | | | | | 0.944 | 0.408 | p<.0001 | | | 0.834 | 0.543 | 0.0216 |
| | | | | | | Saliva Viral Load | -0.19, (-0.269--0.112) | | | p<.0001 | NP Viral Load | -0.061, (-0.136-0.013) | | | 0.107 |
| | | | | | | Days from Symptom Onset | 0.002, (-0.016-0.019) | | | 0.863 | Days from Symptom Onset | 0.015, (-0.002-0.032) | | | 0.089 |

** Log (x10$^6$ cells/ml), * Standard cell counts (x10$^6$ cells/ml)
